# cfDNA concentration as an independent determinant of multi-cancer early detection sensitivity: evidence from a large Indian case-control cohort

**DOI:** 10.64898/2026.07.03.26355665

**Authors:** Swaraj Basu, Prakash Hiremath, Nihesh Rathod, Aditi Chatterjee, Divya Vishwanath, Arunima Ghosh, T. Sthanusubramonian, Shashank Kumar, Krithika Subramanian, R.T. Preetha, Arya Nair, Kannadhasan Sekar, Sarath Ra, Subuhi Yete, G. Bhanumathy, Urvashi Bahadur, Aneesha Radhakrishnan, Abhishek Sarkar, Siddik Uzzaman, Alfiya Beig, Ankita Khan, Badri Padhukasahasram, Lavanya Nemani, Yasodha K. Sivaswamy, Lavanya Bollipalli, Pallavi Ghana, Sameer Phalke, Charles R. Cantor, Sewanti Limaye, Vijay Chandru, Vamsi Veeramachaneni, Ramesh Hariharan

## Abstract

**Background:** The relationship between total cell-free DNA (cfDNA) concentration and multi-cancer early detection (MCED) sensitivity is non-obvious on account of competing considerations. On the one hand, this concentration is elevated in cancer and increases in advanced disease, suggesting higher concentrations may be associated with more biologically active tumors that are easier to detect. On the other hand, this elevation is known to be largely leukocyte-derived, which may dilute tumor-derived DNA (ctDNA) and make detection harder. The net direction of these competing effects on detection sensitivity has not been systematically examined.

**Methods:** EMERGE is an observational case-control study conducted at 43 Indian sites from June 2022–February 2025. It prospectively enrolled and analyzed 1,030 treatment-naïve participants with malignant or benign conditions, most presenting symptomatically, along with 450 controls aged ≥50 years without prior malignancy. Plasma cfDNA underwent targeted hybrid-capture enzymatic methylation sequencing. Classifiers were trained for cancer detection and tissue-of-origin prediction, and tested on the independent validation set. Primary outcomes were the associations between total cfDNA concentration and (i) detection sensitivity and (ii) tissue-of-origin accuracy, evaluated in an independent validation cohort.

**Results:** After adjustment for cancer type, stage, demographic and technical covariates, cfDNA concentration was significantly associated with detection sensitivity (p=6×10^−4^) but not with tissue-of-origin accuracy (p=0.67). At 0.986 specificity (95% CI: 0.968–1.000), stage I sensitivity rose monotonically from 0.52 (95% CI: 0.34–0.69) in the lowest cfDNA concentration tertile to 0.85 (95% CI: 0.73–0.97) in the highest. This association was mechanistically supported by a region-specific increase in hypermethylation scores within regions identified as differentially hypermethylated in TCGA tumor tissue, while panel-wide scores declined. The dissociation between the concentration-sensitivity and concentration–tissue-of-origin associations, together with inverse or insignificant correlations between ctDNA fraction and cfDNA concentration at early stages in published datasets, suggests that the concentration-sensitivity association is partly independent of ctDNA fraction.

**Conclusions:** Total cfDNA concentration is a routinely measured determinant of MCED assay sensitivity, reflecting enrichment of tumor-associated aberrant methylation partly independent of ctDNA fraction—an association likely most pronounced in symptomatic cohorts. Standardized reporting of cfDNA concentration could improve cross-study benchmarking.

**Study Registration:** Clinical Trials Registry, India: CTRI2022/05/042936

**GRAPHICAL ABSTRACT:** 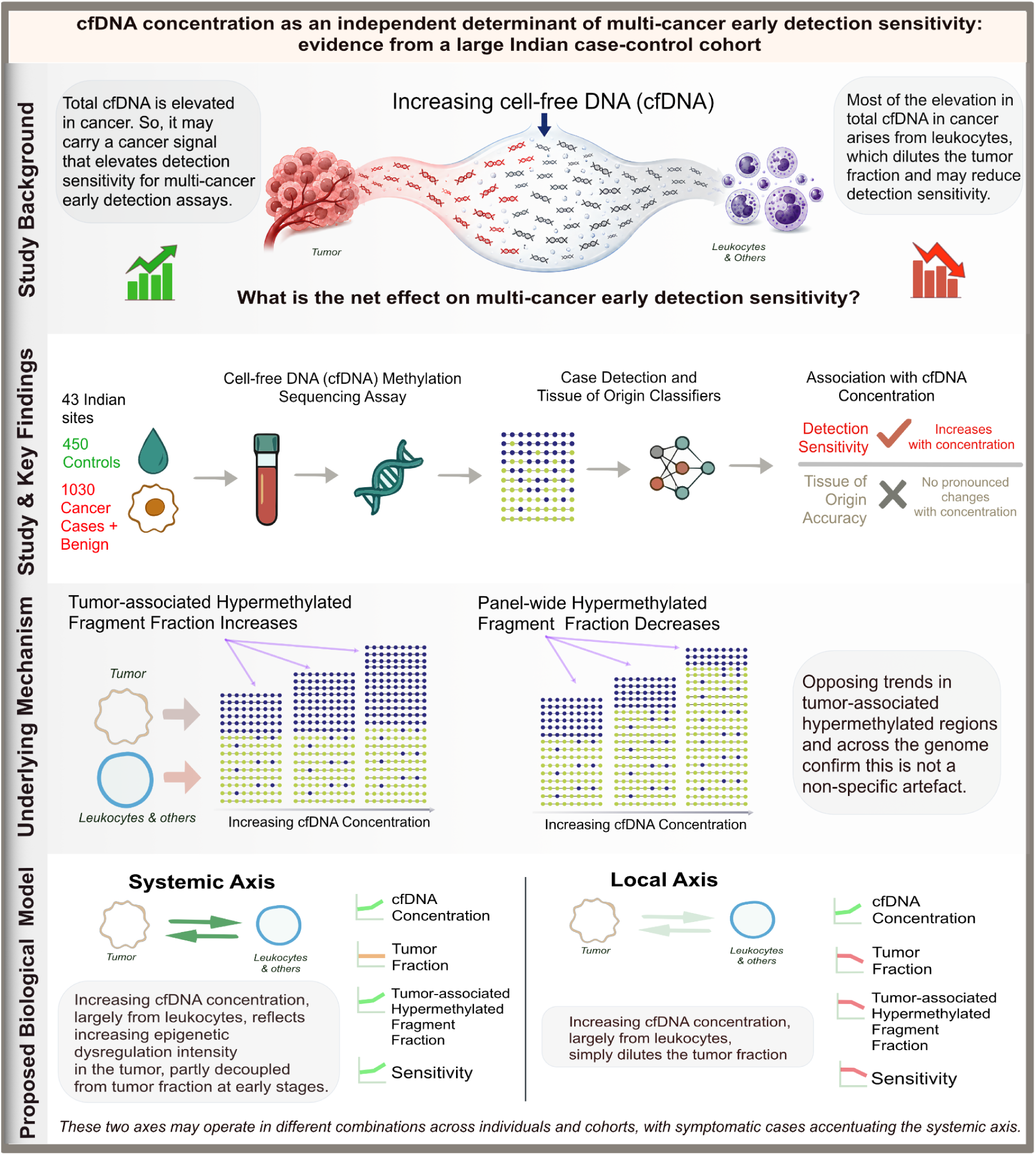

## BACKGROUND

Cell-free DNA (cfDNA)-based multi-cancer early detection (MCED) assays have emerged as a promising approach for detecting malignancies lacking established screening strategies, with tumor-associated methylation and fragmentomic features as leading biomarkers (1–10). Reported stage I sensitivities vary substantially across studies: approximately 15–40% in large Western cohorts (2,11) versus 70–80% in other studies at comparable specificity (3,7). Sensitivity varies by cancer type, disease stage, and ascertainment method, i.e., whether cases were identified through screening or clinical presentation. Additional pre-analytical and biological determinants of assay performance remain incompletely characterized.

The fraction of cfDNA molecules that are tumor-derived, referred to as the circulating tumor DNA (ctDNA) fraction, is regarded as the primary determinant of detection sensitivity (12,13); tumors that shed more DNA into the circulation are easier to detect. Estimating ctDNA fraction requires paired tumor tissue with tumor-derived somatic variants tracked in cfDNA. As a result, ctDNA fraction data are rarely available in MCED studies.

Total cfDNA concentration, by contrast, is routinely measured in all liquid biopsy assays. However, its relationship with detection sensitivity is unclear. Because ctDNA constitutes only a small fraction of total cfDNA, particularly in early-stage disease (14), rising cfDNA concentration is expected to dilute the tumor-derived signal and reduce sensitivity. At the same time, total cfDNA concentration is known to be elevated in cancer (15,16), suggesting it encodes cancer-associated signals that could enhance detection. This apparent paradox is partially resolved by the observation that the excess cfDNA in cancer subjects is primarily leukocyte-derived rather than tumor-derived (16), which again favors a dilution effect. The net relationship between total cfDNA concentration and detection sensitivity is therefore non-obvious and has not been systematically examined.

We hypothesized that total cfDNA concentration is an independent determinant of detection sensitivity and contributes to reported performance differences across cohorts, a question with direct relevance to underlying tumor biology and to cross-study benchmarking. To test this, we applied a targeted enzymatic methylation sequencing assay (17–20) to a large multi-institutional case-control cohort spanning multiple tumor types and disease stages from India, a population substantially underrepresented in the MCED literature. We systematically evaluated cfDNA concentration as a correlate of tumor-associated methylation dysregulation and as a determinant of detection and tissue localization sensitivity. Our findings offer biological insight into the determinants of cfDNA-based detection and suggest practical approaches for improving benchmarking and implementation of methylation-based MCED assays across diverse populations.

## METHODS

### Study Design

This multicenter, observational, case-control study (EMERGE; Clinical Trials Registry, India: CTRI2022/05/042936) prospectively recruited participants across 43 sites in India between June 15, 2022 and February 4, 2025. The protocol was approved by the ethics committees at each participating site (Supplementary Methods 1). All participants provided written informed consent prior to enrollment. The study design is summarized in Fig. 1.

**Figure 1.**
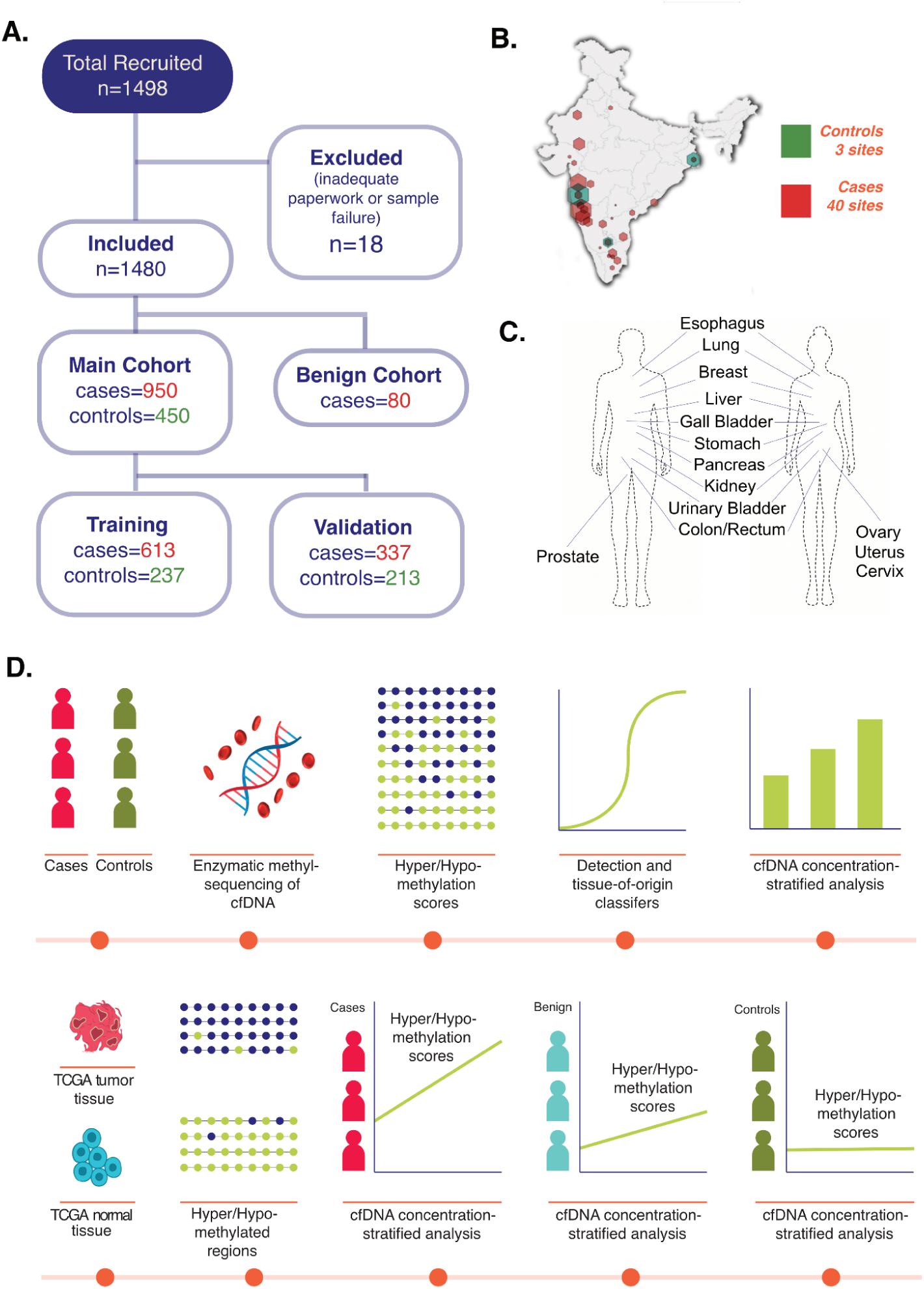
Study design and workflow. **(A) Study Population:** CONSORT diagram of the study cohort. The main cohort is used for classifier training and validation, and the benign cohort is used for further analysis. **(B) Sites:** The distribution of recruitment sites, color coded by cases and controls, with the size of each site being in proportion to the number of samples sequenced. **(C) Cancer Types:** The 14 cancer types in our main cohort. Most samples in the benign cohort also arise from these tissues. **(D) Key Analysis Steps: T**he overall analytical approach, starting with cfDNA enzymatic methyl-sequencing of cohort samples, and using tumor and normal tissue samples from The Cancer Genome Atlas (TCGA) for cross-comparison and to elucidate the underlying mechanism. Both culminate in analysis stratified by cfDNA concentration.

### Participants

Cases included treatment-naïve individuals aged 18 years or older with benign, premalignant, or malignant solid tumors identified through routine clinical care pathways. Most malignant cases were detected upon clinical presentation rather than screen detected. All malignant cases were clinically and/or histologically confirmed and staged according to the AJCC Staging Manual (7th or 8th edition) (21,22). Treating specialists confirmed the diagnosis, tumor type, stage, and treatment-naïve status according to the study criteria. Recruitment monitoring was limited to ensuring representation across cancer types and stages.

The benign cohort comprised individuals presenting with findings suspicious for malignancy who were subsequently confirmed to have benign disease based on standard pathological and/or clinicoradiological assessment. Common diagnoses included inflammatory conditions (e.g., chronic gastritis, colitis, prostatitis, cervicitis), benign cystic lesions (e.g., ovarian cysts, renal cortical cysts, hepatic cysts), benign tumors (e.g., fibroadenoma, leiomyoma, adenomyosis, lipoma, hemangioma), hyperplasia and metaplasia, as well as reactive lymphadenopathy. All cases were negative for dysplasia or malignancy.

The control cohort consisted of asymptomatic individuals aged 50 years or older who underwent clinical evaluation and questionnaire-based assessment by site investigators to exclude prior benign, premalignant, or malignant disease, as well as acute infection. No imaging- or laboratory-based screening was performed. No additional medical exclusions were applied. Control recruitment was monitored to ensure representation of tobacco users and non-users. Pregnant or lactating women were excluded from all cohorts.

### Clinical & Demographic Covariates

Cancer type, stage and tumor size (the maximum reported lesion diameter) were curated from available radiology reports. Tobacco use, alcohol use, gender and age were self-reported by participants and reviewed by the site investigators, while BMI was calculated using height and weight by the site investigators.

### Pre-analytical and Analytical Steps

Pre-analytical and analytical details are in Supplementary Methods 2–4. Peripheral blood was collected in Streck tubes prior to therapy initiation for cases or at enrollment for controls and processed centrally within 72 hours. Plasma isolation, cfDNA extraction, and cfDNA quantification were performed centrally. Quantification was performed on the Agilent 4200 TapeStation with High Sensitivity D1000 ScreenTapes, restricted to fragment lengths 100-1500 to exclude contribution from high molecular weight DNA. This was followed by enzymatic methylation sequencing using a targeted hybrid-capture panel covering approximately 550K genomic regions and 3.98M CpG sites (123 Mb).

Sequencing data were processed to generate alignment, methylation calls, and quality-control metrics (Supplementary Methods 5).

One of the challenges of methylation assays is incomplete conversion of unmethylated cytosines. This can appear as methylation and compromise specificity. Completeness of conversion was assessed using non-CpG cytosines (expected to be unmethylated). Fragments failing predefined conversion thresholds were excluded at the fragment level rather than rejecting entire samples with inadequate non-CpG cytosine conversion; we refer to this as **the rescue step** (Supplementary Methods 6).

Fragment-level methylation patterns were analyzed to compute the fraction of fragments in each panel region with ≥80% of CpGs methylated and additionally ≥3 CpGs mapping to the region (**Ge80**), and the analogous fraction with <20% of CpGs methylated (**Lt20).** (Supplementary Methods 7). These scores were used for case-control classifier construction (Supplementary Methods 9–13). Additional fragmentomic features (fragment size, end-motif frequency, and nucleosome protection; Supplementary Methods 8) were computed for tissue-of-origin classification (Supplementary Methods 16).

The main cohort was partitioned into training and independent validation cohorts via stratified random sampling by cancer type, stage, age, and gender. Classifier training thresholds were locked prior to application on the independent validation set.

### Outcomes

The primary outcomes were the association between cfDNA concentration and cancer detection sensitivity, and the association between cfDNA concentration and tissue-of-origin prediction accuracy, evaluated in the independent validation cohort.

### Sample Size

Sample size was not pre-specified by formal power or precision calculations. However, the achieved validation cohort sizes provided adequate precision (with >186 controls, the one-sided 95% lower confidence bound for specificity exceeded 99% minus 1.2%, and with >87 stage I/II cancer cases each, the lower bound for early-stage sensitivity exceeded 70% minus 8.1%).

### Consistency Analysis of Methylation Scores

To assess consistency of methylation scores in our cfDNA data with those in orthogonal datasets, differentially methylated regions identified in TCGA tumor tissues were evaluated in our cfDNA dataset, and cfDNA-derived classifier regions were independently evaluated against TCGA tumor tissue methylation profiles (Supplementary Methods 17). Enrichment analysis of classifier regions was performed to determine functional significance (Supplementary Methods 22).

### Classifier Performance and Robustness

Sensitivity and specificity for discriminating between cases and controls in cross-validation and independent validation were reported to characterize overall assay performance, at a pre-specified training specificity threshold. Confidence intervals were estimated using bootstrap resampling (Supplementary Methods 14, 16).

Tissue-of-origin (TOO) performance was assessed using top-1 and top-2 accuracy metrics in both cross-validation and independent validation, after grouping the 14 cancer types studied into 9 anatomical tissue groups. Top-1 accuracy was defined as the proportion of samples for which the predicted tissue of origin matched the true anatomical group, whereas top-2 accuracy was defined as the proportion for which the true anatomical group was included among the two highest-ranked predicted tissues of origin.

To evaluate conversion efficiency and rescue-step performance, sensitivity, specificity and fraction of reads dropped were assessed upon omission of this step (Supplementary Methods 15).

To evaluate robustness to spatial, temporal, and technical drift, including drift on account of reagent lots, the main cohort was repartitioned into non-overlapping training and independent validation sets using site-wise, collection time-wise, and sequencing batch-wise splits. In each case, classifiers were developed on the training set and thresholds were locked before independent validation was conducted on the held-out validation set. (Supplementary Methods 20). To assess robustness to measured confounding, the association between classifier score and cancer status was evaluated using unadjusted and adjusted logistic regression models, with adjustment for demographic, lifestyle, and technical covariates. To assess sensitivity to covariate imbalance, inverse probability weighting was applied and weighted and unweighted estimates were compared for stability. Stratified analyses by tobacco usage status and by control recruitment site were performed to assess the potential impact of tobacco enrichment in controls and geographic concentration of control sites respectively (Supplementary Methods 21).

### Association Analysis Between Classifier Sensitivity and cfDNA Concentration

Classifier sensitivity was evaluated as a function of tumor size and cfDNA concentration using rolling-window estimates stratified by disease stage.

To assess whether the association between cfDNA concentration and detection sensitivity was independent of cancer type and stage and other key covariates, multivariable logistic regression models (using statsmodels GLM in Python) were fitted with detection status as the binary outcome, adjusted for various demographic, clinical and technical covariates. Non-linear effects of cfDNA concentration were modelled using natural cubic splines, and the statistical significance of the concentration association was assessed using a joint Wald test of all spline terms. (Supplementary Methods 19). A similar analysis was performed to evaluate the association between cfDNA concentration and tissue-of-origin top-1 and top-2 accuracy.

To provide a mechanistic explanation for the cfDNA concentration-sensitivity association, samples were stratified into tertiles of cfDNA concentration within each stage and in the benign and control groups. Ge80 and Lt20 scores were averaged over three region sets: regions identified as hypermethylated in TCGA tumor tissues relative to adjacent normals (Supplementary Methods 17), regions identified as hypomethylated in TCGA tumor tissues relative to adjacent normals, and across all 550K panel regions. Mean scores and 95% Wald confidence intervals were computed within each concentration tertile and stage stratum and plotted to assess directional trends.

### Relationship Between cfDNA Concentration and Circulating Tumor DNA (ctDNA) Fraction

Because ctDNA fraction was not directly measured in our study cohort, we analysed published treatment-naïve cancer datasets in which both total cfDNA concentration and mutation-derived estimates of ctDNA fraction were available. In each dataset, ctDNA fraction was estimated from circulating mutant allele frequencies (cMAF) of confirmed somatic mutations identified through matched tumor sequencing. Within each stage, the association between cfDNA concentration and cMAF-estimated ctDNA fraction was assessed using Spearman rank correlation. Analyses were performed separately by stage because both cfDNA concentration and ctDNA fraction are strongly stage-dependent.

### Benchmarking Across Studies

To compare stage-wise performance across studies with differing cancer-type compositions, stage-specific sensitivities were standardized using binomial generalized linear models incorporating cancer type, study, stage, and study-stage interaction terms. Marginal sensitivities were averaged over the pooled cancer-type distribution, and uncertainty was estimated using nonparametric bootstrap resampling (Supplementary Methods 18).

## RESULTS

### Cohort Characteristics and Study Design

A total of 1,480 participants recruited across 43 sites were included in this analysis. The main cohort comprised 950 cancer cases spanning 14 cancer types across major organ systems, and 450 controls, randomly partitioned into training and independent validation sets. An additional cohort with 80 benign cases comprising lesions arising largely in organs represented among the 14 cancer types in the main cohort was used to study classifier behavior (Fig. 1A-C).

Supplementary Fig. 1 outlines the pre-analytical and analytical steps. Briefly, blood was collected in Streck tubes and received at a central laboratory within 72 hours, where plasma was extracted and stored. cfDNA was extracted from retrieved plasma and quantified. Extracted cfDNA underwent targeted enzymatic methyl-sequencing, followed by secondary analysis and methylation calling. A fragment-level rescue strategy was applied prior to feature computation to mitigate the effects of incomplete conversion. Region-level methylation and fragmentomic features were then computed and used for classifier training with a pre-specified specificity threshold lock. Classifier performance was evaluated in the independent validation cohort, with additional robustness and confounder analyses performed across technical, demographic, and temporal variables. The association of classifier behaviour with cfDNA concentration on the validation and benign cohorts was assessed as detailed in Methods (Fig. 1D).

Baseline characteristics of the main cohort are summarized in Table 1 (per-sample details in Supplementary Table 1). Cases and controls were broadly comparable in age (60.0 vs 61.0 years), BMI (23.4 vs 22.9 kg/m^2^), and sequencing quality metrics including average coverage (70.1 vs 70.2×), fragment counts (31.1 million in both), and fraction of reads lost on rescue (0.13 in both). Controls had a higher proportion of males (57.3% vs 47.1%) and markedly greater prevalence of tobacco use (69.8% vs 22.3%) and alcohol use (20.0% vs 14.3%). Cancer types were diverse, with colorectal, stomach, breast, and lung cancers being most frequent; stage distribution was balanced across stages I–IV (27.3%, 26.7%, 23.7%, 22.3%), and median tumor size increased with stage (3.0, 4.4, 6.0, and 6.0 cm for stages I–IV respectively), with measurements unavailable for 334 of 950 cases. As expected, cfDNA concentration was substantially higher in cases than controls (8.46 vs 4.21 ng/mL). 20 ng of cfDNA was taken for most samples, with only 273 of 1400 samples using lower input amounts. Controls had shorter plasma storage duration prior to cfDNA extraction (71 vs 218 days) and were less frequently sequenced on NovaSeq X+ relative to NovaSeq 6000 (69.1% vs 97.4% in cases). Key variables above were included as covariates in multivariable sensitivity analyses. Distributions of all characteristics were similar across training and validation cohorts.

**Table 1.** Main Cohort Characteristics. Key demographic, clinical, pre-analytical, analytical and sequencing covariates are summarized. Missing value counts are shown in curly brackets { }, interquartile ranges or IQRs in square brackets []. Medians and IQRs are shown for continuous parameters, and counts and percentages for categorical parameters, with percentages shown in round brackets.

|  | Cases | Controls | Training Cases | Training Controls | Validation Cases | Validation Controls |
| --- | --- | --- | --- | --- | --- | --- |
| Total N | 950 | 450 | 613 | 237 | 337 | 213 |
| **DEMOGRAPHIC** |  |  |  |  |  |  |
| Age (years) | 60.0 [50.0–68.8] {0} | 61.0 [55.0–70.0] {0} | 59.0 [49.0–68.0] {0} | 60.0 [55.0–68.0] {0} | 60.0 [51.0–69.0] {0} | 62.0 [55.0–72.0] {0} |
| BMI (kg/m<sup>2</sup>) | 23.4 [21.3–25.9] {1} | 22.9 [20.7–26.0] {0} | 23.4 [21.3–25.9] {1} | 22.7 [20.8–26.0] {0} | 23.3 [21.3–25.8] {0} | 23.2 [20.7–26.1] {0} |
| Gender Male | 447 (47.1%) {0} | 258 (57.3%) {0} | 286 (46.7%) {0} | 145 (61.2%) {0} | 161 (47.8%) {0} | 113 (53.1%) {0} |
| Tobacco User | 212 (22.3%) {0} | 314 (69.8%) {0} | 141 (23.0%) {0} | 149 (62.9%) {0} | 71 (21.1%) {0} | 165 (77.5%) {0} |
| Alcohol User | 136 (14.3%) {16} | 90 (20.0%) {1} | 83 (13.5%) {12} | 41 (17.3%) {1} | 53 (15.7%) {4} | 49 (23.0%) {0} |
| **CLINICAL** |  |  |  |  |  |  |
| Stage I | 259 (27.3%) | - | 160 (26.1%) | - | 99 (29.4%) | - |
| Stage II | 254 (26.7%) | - | 167 (27.2%) | - | 87 (25.8%) | - |
| Stage III | 225 (23.7%) | - | 146 (23.8%) | - | 79 (23.4%) | - |
| Stage IV | 212 (22.3%) | - | 140 (22.8%) | - | 72 (21.4%) | - |
| Tumor size Stage I (cm) | 3.0 [1.5–5.0] {109} | - | 3.0 [1.5–5.4] {66} | - | 2.8 [1.5–4.6] {43} | - |
| Tumor size Stage II (cm) | 4.4 [2.7–6.5] {82} | - | 4.2 [2.7–6.3] {56} | - | 5.0 [3.0–6.5] {26} | - |
| Tumor size Stage III (cm) | 6.0 [4.5–8.2] {65} | - | 6.1 [4.8–8.4] {40} | - | 5.5 [4.0–8.0] {25} | - |
| Tumor size Stage IV (cm) | 6.0 [4.5–9.0] {78} | - | 6.0 [4.3–8.4] {50} | - | 7.2 [4.7–9.6] {28} | - |
| Cancer: Breast | 88 (9.3%) | - | 59 (9.6%) | - | 29 (8.6%) | - |
| Cancer: Cervix | 71 (7.5%) | - | 43 (7.0%) | - | 28 (8.3%) | - |
| Cancer: Colorectal | 173 (18.2%) | - | 114 (18.6%) | - | 59 (17.5%) | - |
| Cancer: Esophagus | 62 (6.5%) | - | 39 (6.4%) | - | 23 (6.8%) | - |
| Cancer: Gall Bladder | 43 (4.5%) | - | 28 (4.6%) | - | 15 (4.5%) | - |
| Cancer: Kidney | 43 (4.5%) | - | 30 (4.9%) | - | 13 (3.9%) | - |
| Cancer: Liver | 27 (2.8%) | - | 16 (2.6%) | - | 11 (3.3%) | - |
| Cancer: Lung | 85 (8.9%) | - | 48 (7.8%) | - | 37 (11.0%) | - |
| Cancer: Ovary | 77 (8.1%) | - | 47 (7.7%) | - | 30 (8.9%) | - |
| Cancer: Pancreas | 62 (6.5%) | - | 40 (6.5%) | - | 22 (6.5%) | - |
| Cancer: Prostate | 47 (4.9%) | - | 34 (5.5%) | - | 13 (3.9%) | - |
| Cancer: Stomach | 93 (9.8%) | - | 56 (9.1%) | - | 37 (11.0%) | - |
| Cancer: Urinary Bladder | 41 (4.3%) | - | 31 (5.1%) | - | 10 (3.0%) | - |
| Cancer: Uterus | 38 (4.0%) | - | 28 (4.6%) | - | 10 (3.0%) | - |
| **(PRE &) ANALYTICAL** |  |  |  |  |  |  |
| Plasma Storage Time (days) | 218.0 [113.0–340.0] {0} | 71.0 [58.0–151.0] {0} | 224.0 [120.0–362.0] {0} | 69.0 [55.0–132.0] {0} | 206.0 [103.0–317.0] {0} | 110.0 [60.0–237.0] {0} |
| cfDNA Yield (ng/mL plasma) | 8.46 [4.52–19.93] {4} | 4.21 [2.89–6.22] {0} | 8.47 [4.54–19.58] {1} | 4.11 [2.94–6.03] {0} | 8.36 [4.49–20.31] {3} | 4.35 [2.80–6.59] {0} |
| cfDNA Input (ng) | 20.0 [20.0–20.0] {0} | 20.0 [16.8–20.0] {0} | 20.0 [20.0–20.0] {0} | 20.0 [17.6–20.0] {0} | 20.0 [20.0–20.0] {0} | 20.0 [16.4–20.0] {0} |
| Library Yield (ng) | 907.6 [677.0–1123.5] {0} | 773.0 [552.6–1030.8] {0} | 910.8 [680.0–1140.0] {0} | 786.6 [549.0–1016.6] {0} | 884.0 [673.2–1104.0] {0} | 764.0 [554.4–1052.0] {0} |
| Sequencer (NovaSeq X+/6000) | 925 (97.4%) {0} | 311 (69.1%) {0} | 597 (97.4%) {0} | 153 (64.6%) {0} | 328 (97.3%) {0} | 158 (74.2%) {0} |
| **SEQUENCING** |  |  |  |  |  |  |
| Average Coverage | 70.1 [59.1–89.0] {0} | 70.2 [59.1–82.9] {0} | 72.6 [59.5–92.2] {0} | 70.0 [59.2–80.9] {0} | 66.8 [58.4–82.4] {0} | 70.3 [58.8–84.9] {0} |
| NonCpG C Conversion <99% | 194 (20.4%) {0} | 99 (22.0%) {0} | 135 (22.0%) {0} | 59 (24.9%) {0} | 59 (17.5%) {0} | 40 (18.8%) {0} |
| No. of Fragments (million) | 31.1 [25.5–42.8] {0} | 31.1 [25.6–37.2] {0} | 32.1 [26.3–45.9] {0} | 31.1 [25.3–37.2] {0} | 29.8 [25.0–39.3] {0} | 31.5 [26.0–36.9] {0} |
| Fraction Reads Lost on Rescue | 0.13 [0.12–0.16] {0} | 0.13 [0.11–0.17] {0} | 0.13 [0.12–0.16] {0} | 0.13 [0.11–0.17] {0} | 0.13 [0.12–0.16] {0} | 0.13 [0.11–0.16] {0} |

### Pan-Cancer Hypermethylation Patterns Underlie Classifier Detection

Before assessing the association between cfDNA concentration and classifier sensitivity, we established the baseline characteristics of the classifier and the epigenetic basis for its discriminative ability.

The classifier achieved an AUC of 0.960 (95% CI 0.947–0.973) in independent validation. At the pre-specified 99% training specificity threshold, validation specificity was 0.986 (95% CI 0.968–1) and overall sensitivity was 0.72, with stage I sensitivity of 0.72 (95% CI 0.63–0.80) rising to 0.83 at stage IV. The benign cohort detection rate was 0.33 (95% CI 0.24–0.44). TOO top-1 accuracy was 0.76 (95% CI 0.71–0.80) in independent validation. Full performance characteristics, robustness analyses, and TOO results are reported in Supplementary Results 1 and Supplementary Fig. 2–3. cfDNA concentration was not an input variable for the classifier and hence represents an independent measurement.

The detection classifier used 4,384 cfDNA-derived regions identified in training, covering 149,242 CpGs with a total sequence length of ∼3.23 Mb and a median raw overlapping read count of ∼4M in the main cohort. To establish the biological basis of the classifier, we cross-referenced these regions against tissue methylation trends in The Cancer Genome Atlas (TCGA) (Supplementary Table 2). While panel-wide TCGA tumor tissues showed expected global hypomethylation relative to adjacent normal/healthy tissues (Fig. 2B), as observed in prior reports (23), these specific 4,384 regions demonstrated the inverse trend, showing tumor-specific hypermethylation across all 13 evaluated cancer types (Fig. 2A), and suggesting that these pan-cancer hypermethylation patterns underlie classifier detection.

**Figure 2.**
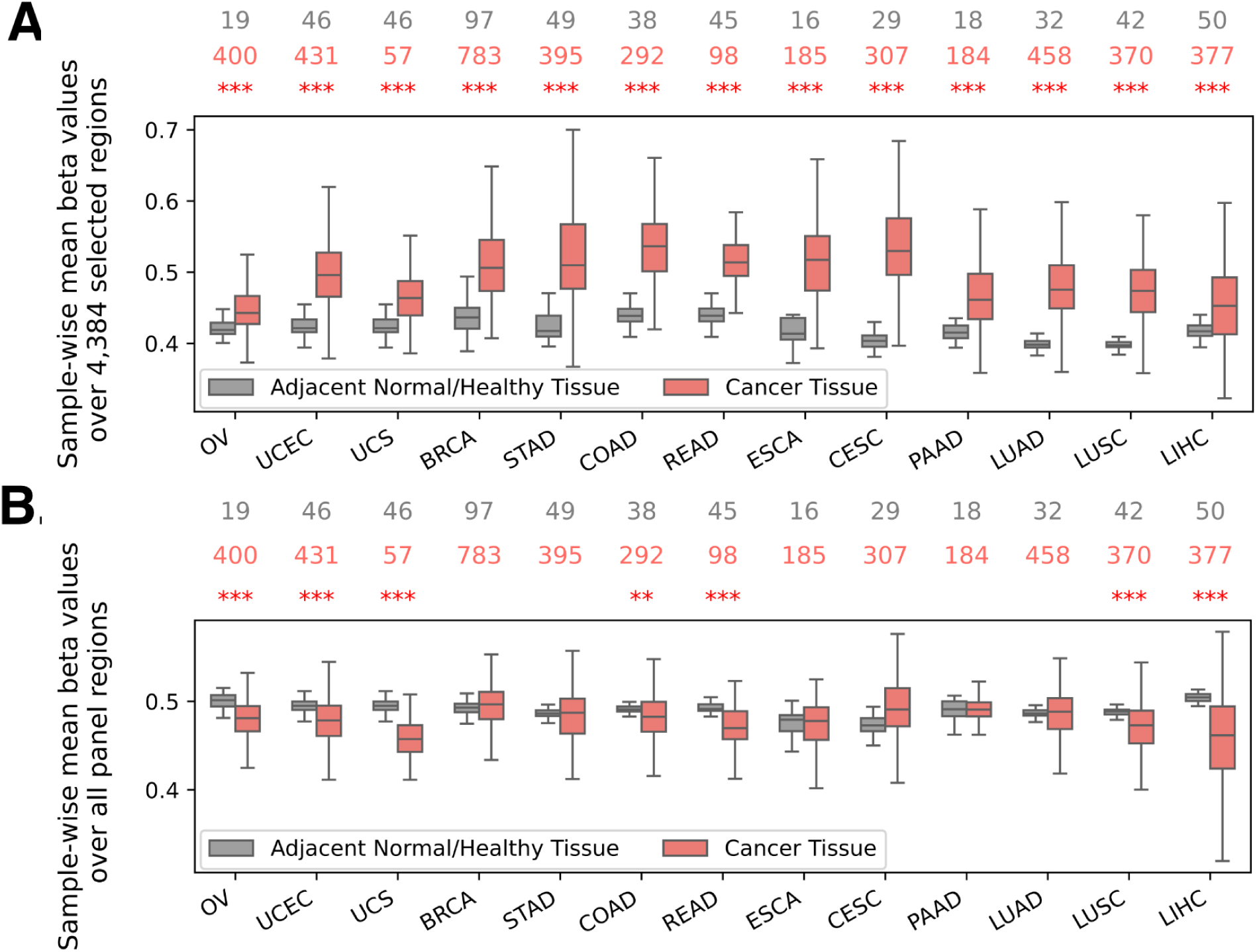
Discriminatory basis of classifier regions in TCGA tumor tissue samples. (**A)** Average beta value distributions for probes overlapping the 4,384 hypermethylated regions used by our classifier, in TCGA cancer samples and in adjacent normal/healthy tissue are shown, for 13 cancer types. Sample counts of each type are indicated at the top in the corresponding color. Q-values assessed by Mann-Whitney one-sided test for relative **hypermethylation** in cancer tissue are also shown, in conventional star notation. (**B)** Average beta value distributions for **all probes** in TCGA cancer samples and in adjacent normal/healthy tissue are shown, for 13 cancer types. Sample counts are again indicated as above. Q-values assessed by Mann-Whitney one-sided test for relative **hypomethylation** in cancer tissue are also shown, in conventional star notation. Cancer types: OV: Ovarian Serous Cystadenocarcinoma, UCEC: Uterine Corpus Endometrial Carcinoma, UCS: Uterine Carcinosarcoma, BRCA: Breast Invasive Carcinoma, STAD: Stomach Adenocarcinoma, COAD: Colon Adenocarcinoma, READ: Rectum Adenocarcinoma, ESCA: Esophageal Carcinoma, CESC: Cervical Squamous Cell Carcinoma and Endocervical Adenocarcinoma, PAAD: Pancreatic Adenocarcinoma, LUAD: Lung Adenocarcinoma, LUSC: Lung Squamous Cell Carcinoma, LIHC: Liver Hepatocellular Carcinoma.

Functional enrichment analyses of the 4,384 regions suggested enrichment for CpG-dense promoter loci and implicated conserved Polycomb-associated developmental programs and repressive chromatin states (H3K27me3/H3K9me3), without strong tissue-specific enrichment, consistent with broadly shared pan-cancer epigenetic repression biology (Supplementary Results 4).

### cfDNA Concentration Independently Predicts Detection Sensitivity

We next studied how sensitivity varied with cfDNA concentration. Rolling window analyses confirmed that the fraction of samples with cfDNA concentration exceeding 3.7 ng/mL visually tracked classifier sensitivity better than tumor size did (Fig. 3A). It also demonstrated a concentration-sensitivity gradient present even in benign disease but absent in healthy controls (Fig. 3B).

**Figure 3.**
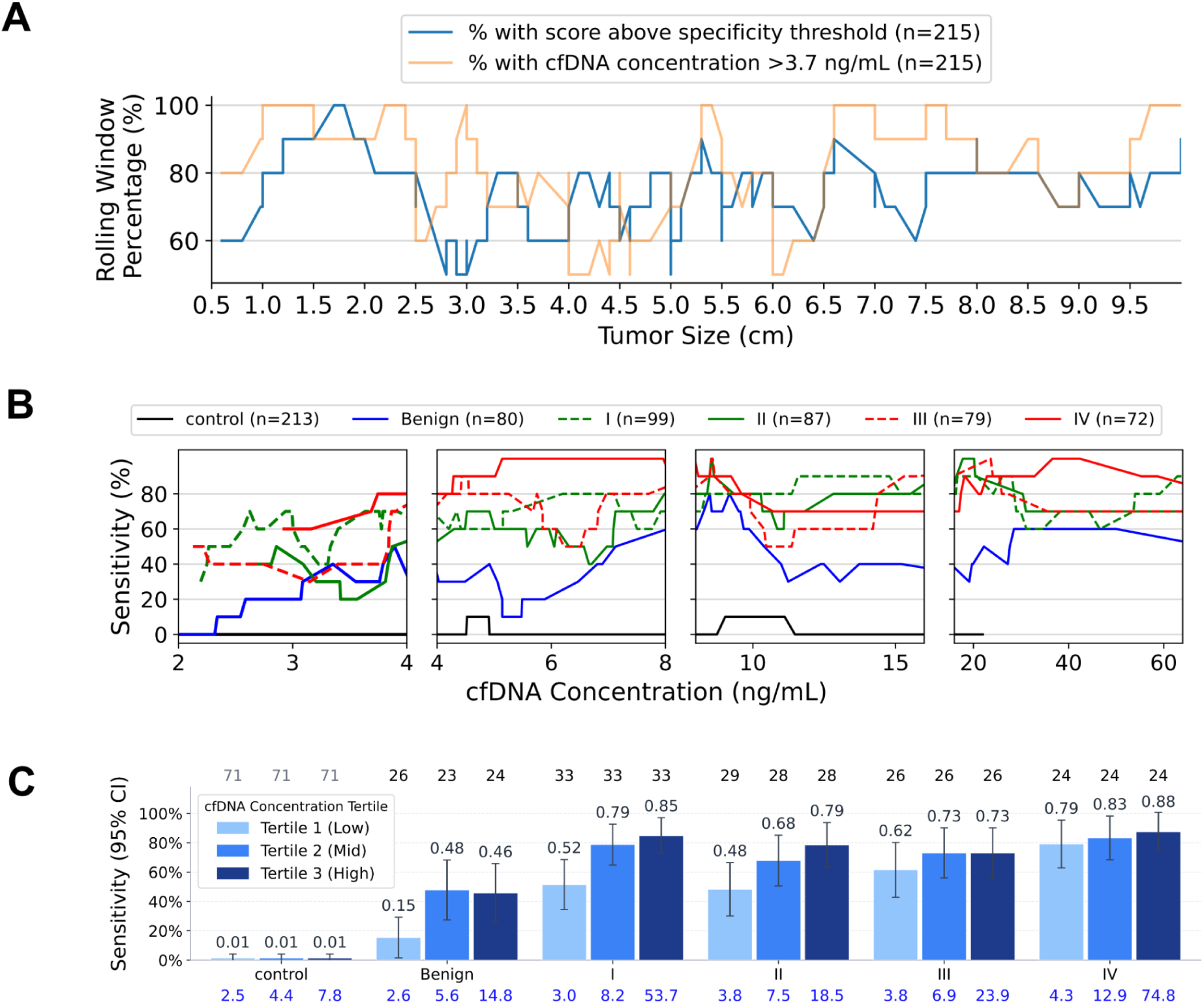
Association between cfDNA concentration and classifier sensitivity in the independent validation and benign cohorts. **(A)** Classifier sensitivity in rolling windows of 10 samples after sorting samples by tumor size is shown as a profile plot in blue. The fraction of samples in the same rolling windows with cfDNA > 3.7 ng/mL is shown in peach. Tumor size was available for only 215/337 samples in the independent validation set. **(B)** Classifier sensitivity in rolling windows of 10 samples after sorting samples by cfDNA concentration, and stratified by disease stage, is shown as profile plots. Different x-axis scales are used in different ranges for cfDNA concentration. The number of samples in each disease stage is indicated in the legend. **(C)** Classifier sensitivity with 95% CI (Wald) for concentration tertiles within each stage, shown as bar plots. The number of samples in each tertile within each stage is indicated above the plot. The median concentration value for each tertile-stage combination is indicated in blue below the bar.

Within most stages, detection rates increased monotonically across concentration tertiles (Fig. 3C): detection rates in the symptomatic benign cohort rose from 0.15 in the lowest tertile to 0.46 in the highest, stage I rose from 0.52 in the lowest tertile to 0.85 in the highest; stage IV from 0.79 to 0.88; controls remained near-zero across all tertiles. Together, these findings suggest that cfDNA concentration is associated with detection sensitivity.

To confirm that this relationship was not driven by differences in cancer type or stage distribution across concentration strata, we fitted a multivariable logistic regression model adjusting for both. The overall effect of cfDNA concentration on sensitivity was statistically significant (joint test of spline terms: χ^2^=24.9, df=4, p=5.2×10^−5^). Statistical significance remained upon further adjustment for age, gender, BMI, tobacco usage, alcohol usage, plasma storage time, and sequencer (χ^2^=22.6, df=4, p=1.5×10^−4^).

To assess if the cfDNA concentration-sensitivity association was on account of the number of independent fragments being sequenced, we note that 20 ng of extracted cfDNA was used for ∼80% of samples, and smaller amounts were used for the remainder; adjustment for the actual assay input amount retained significance at similar levels (χ^2^=21.2, df=4, p=2.9×10^−4^). Further adjustment for average read coverage after duplicate removal also retained significance (χ^2^=19.5, df=4, p=6.1×10^−4^).

cfDNA concentration has been reported to exhibit circadian variation, with morning samples showing approximately 63% higher concentrations than afternoon and evening samples (24). To assess whether this could confound the concentration-sensitivity association, collection times were categorized into three-hour windows and included as a covariate; the association still remained significant (χ^2^=19.6, df=4, p=6.0×10^−4^).

Among the covariates, only stage (p=7.8×10^−4^), age (p=3×10^−3^) and average coverage (p=3.3×10^−2^) also reached statistical significance. Stage is a well-established determinant of sensitivity. The age association was directionally consistent with greater epigenetic dysregulation in older individuals: sensitivity increased from 0.63 in the 40–50 age bracket to 0.85 in the 70–80 bracket, with intermediate values of 0.73 and 0.71 in the 50–60 and 60–70 brackets respectively. Mean coverage showed a small inverse association with sensitivity (0.71 in the 40×–50× range versus 0.68 in the 80×–90× range); higher coverage may partially reflect preferential flow cell binding of shorter cfDNA fragments—a known Illumina sequencing artifact—which are less likely to carry the longer hypermethylated reads that contribute to classifier detection; however, this explanation remains inferential and the effect size is small. Importantly, cfDNA concentration significance was not attenuated by stage, age, or coverage adjustment, indicating that these capture partly independent sources of sensitivity variation.

Together, these analyses establish that cfDNA concentration is an independent predictor of detection sensitivity, robust to cancer type, stage, demographic, clinical and technical variation, motivating investigation of the biological mechanism underlying this association.

### Tumor-Associated Aberrant Methylation Underlies the cfDNA Concentration-Sensitivity Association

To establish the mechanism underlying this association, we derived tumor-specific aberrantly methylated regions from independent tissue datasets and assessed the methylation status of these regions in our main cohort. Methylation status in our cohort was reflected by the Ge80 and Lt20 scores, the former representing the fraction of hypermethylated fragments, and the latter representing the fraction of hypomethylated fragments. We studied how these scores, when aggregated over the chosen regions, varied with cfDNA concentration.

Analysis of TCGA tumor tissue relative to adjacent normal/healthy tissue (Supplementary Table 6) identified 10,225 regions as hypermethylated in tumor tissue, and 14,553 regions as hypomethylated.

In the 10,225 hypermethylated regions, aggregate cfDNA Ge80 hypermethylation scores in the main cohort increased progressively along a biological continuum from healthy controls to symptomatic benign conditions, early-stage, and late-stage malignancies (Fig. 4A–C).

**Figure 4.**
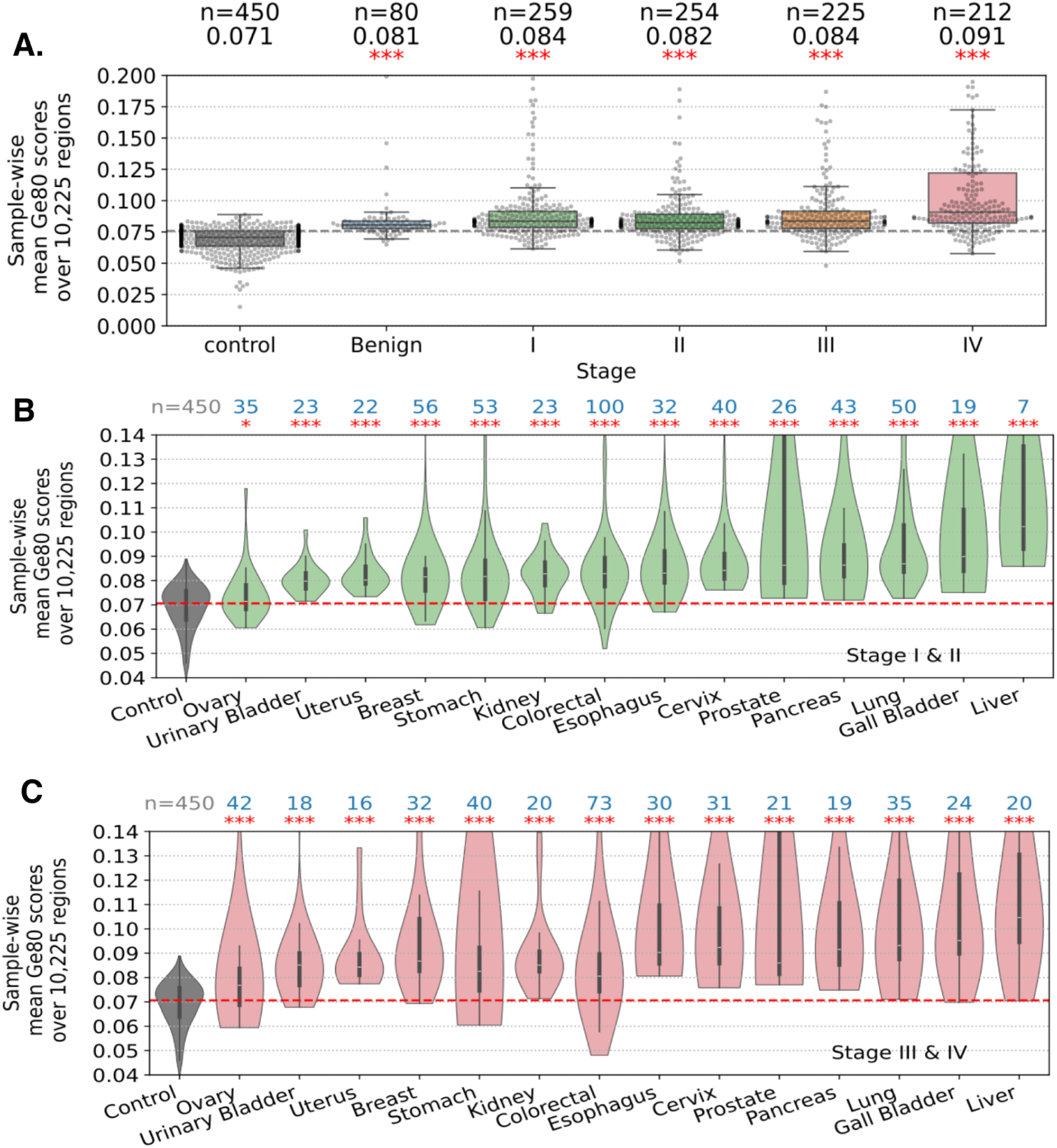
cfDNA methylation status (in the main cohort) of 10,225 regions hypermethylated in TCGA tumor tissue. **(A)** Ge80 scores (the fraction of hypermethylated fragments) averaged over regions in the Twist Human Methylome panel that overlap the chosen 10,225 regions shown as box plots, stratified by disease stage. The horizontal dashed line is the 75th percentile of the control values. Median values and counts in each group are shown above the plot. Q values obtained by one-sided Mann-Whitney testing for hypermethylation in each group relative to controls are shown in standard star notation. **(B)** The same as above but stratified by cancer type instead, for stage I and II samples, and shown as violin plots. Counts of each cancer type are shown above. Q values obtained by one-sided Mann-Whitney testing for hypermethylation in cancer types relative to controls are shown in standard star notation. The dashed line indicates the median control value. **(C)** Same as above, but for stage III and IV samples.

We further characterized this progression by stratifying samples into within-stage cfDNA concentration tertiles. Ge80 scores increased alongside cfDNA concentration tertiles for most stages, while remaining flat in healthy controls (Fig. 5A). Conversely, panel-wide Ge80 scores decreased with increasing cfDNA concentration across stages (Fig. 5B), reflecting the global cancer-associated hypomethylation observed in prior reports (23) and also noted in Fig. 2B. This opposite behavior of region-specific and panel-wide patterns confirms the selective association of cfDNA concentration with tumor-associated hypermethylation signal and argues against a nonspecific effect of cfDNA concentration. Consistent patterns occurred in the 14,553 TCGA-hypomethylated regions: Lt20 scores increased slightly with concentration across stages (Fig. 5C), while panel-wide Lt20 scores also rose slightly (Fig. 5D), reflecting global cancer-associated hypomethylation as expected.

**Figure 5.**
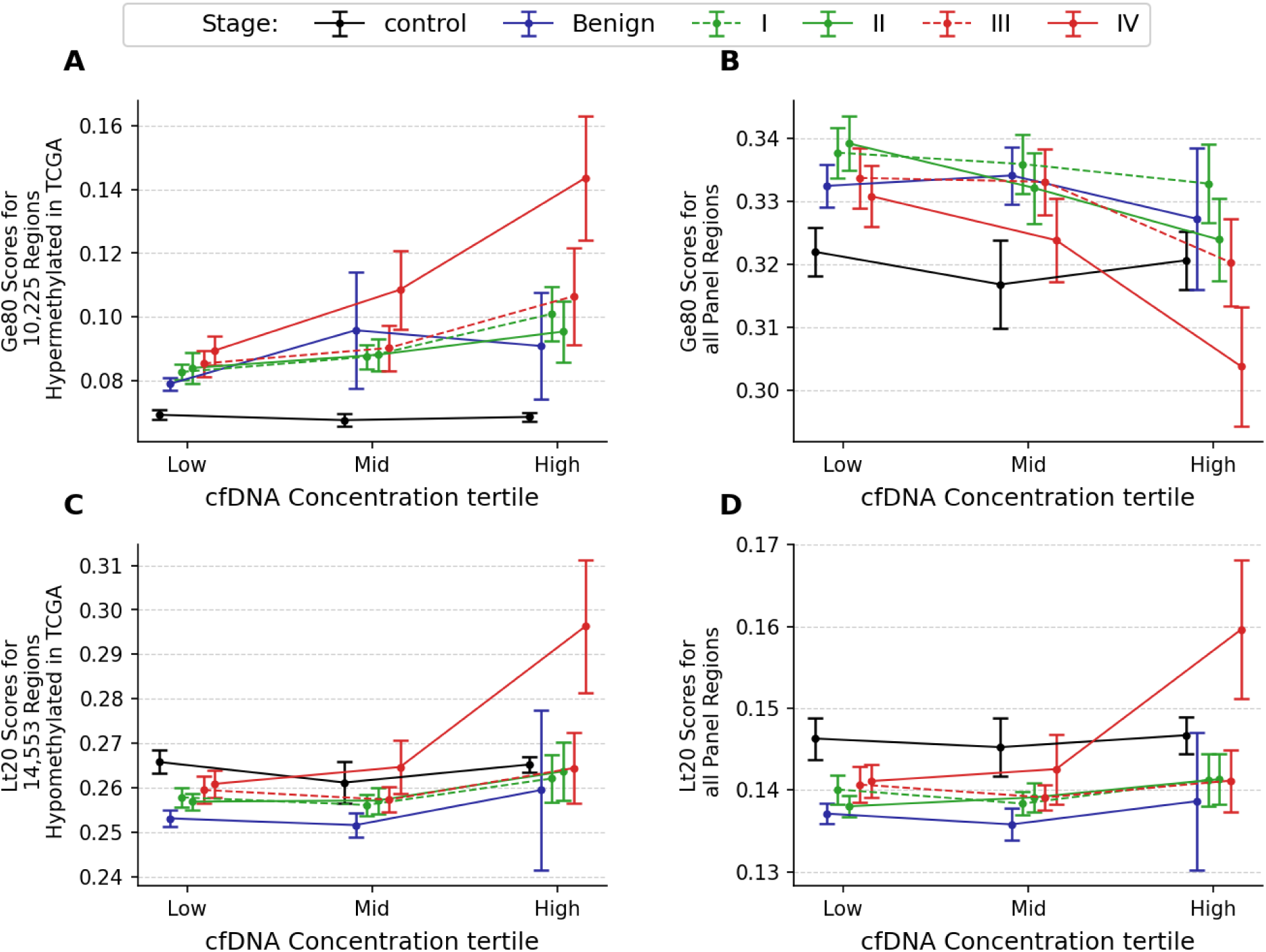
Variation of hypermethylation and hypomethylation scores with cfDNA concentration in the main cohort. Each plot shows the mean and 95% CI (Wald) for a chosen metric, stratified by stage-concentration tertile combinations. Concentration tertiles are defined within each stage. The metrics in the depiction are: **(A)** Mean cfDNA Ge80 hypermethylation scores for 10,225 hypermethylated regions identified from TCGA analysis. **(B)** Mean cfDNA Ge80 hypermethylation scores for all 550K panel regions. **(C)** Mean cfDNA Lt20 hypomethylation scores for 14,553 hypomethylated regions identified from TCGA analysis. **(D)** Mean cfDNA Lt20 hypomethylation scores for all 550K panel regions.

These concentration-dependent trends were robust to alternative stratification by tumor size or absolute concentration bins (Supplementary Figs. 4–6, respectively).

In multivariable ordinary least squares regression adjusting for cancer type and stage, the independent effect of cfDNA concentration on both hypermethylation and hypomethylation scores remained highly significant (p<0.001), confirming that total cfDNA concentration is strongly associated with tumor-associated region-specific aberrant methylation signal. In particular, hypermethylation patterns underlie classifier detection as noted earlier and the strong enrichment of hypermethylated fragments (Fig. 5A) is consistent with improved classifier sensitivity.

### Evidence Suggests Early-Stage Concentration–Sensitivity Associations are Not Fully Explained by ctDNA Fraction

We next asked whether the cfDNA concentration-dependent gain in detection sensitivity reflects an overall increase in ctDNA fraction. An increasing fraction would explain the rise in tumor-associated aberrant methylation signal. Conversely, an unchanging or decreasing fraction would suggest that rising cfDNA enriches tumor-associated aberrant methylation signal via non-tumor-derived pathways.

Because paired tumor tissue was not available in this study, ctDNA fraction was not directly measured in our cohort. We therefore examined the relationship between ctDNA fraction—estimated by circulating mutant allele frequency (cMAF)—and total cfDNA concentration in three independent published datasets where both quantities were available. These datasets (Supplementary Table 7) differed in cancer type and stage distribution, case ascertainment method, and sample size, but shared treatment-naïve status and mutation-based cMAF estimation from matched tissue sequencing, enabling a directional cross-cohort assessment. These analyses are indirect and should be interpreted accordingly.

In the Cohen et al. dataset (11), comprising 1,005 clinically detected cases spanning eight cancer types, cMAF data were available for 138 cases with stomach, colorectal, liver or esophageal cancer. Analysis of these 138 cases showed non-significant inverse correlation in stages II and III (ρ=−0.08 and ρ=−0.11 respectively) between cfDNA concentration and ctDNA fraction; stage I was unevaluable (n=5) and stage IV had no observations (Fig. 6B). Notably, across all 1,005 cases, cfDNA concentration was significantly associated with classifier sensitivity after adjusting for cancer type and stage (χ^2^=60.5, df=4, p=2.18×10^-12^), consistent with the findings in our cohort (Supplementary Fig. 7B).

**Figure 6.**
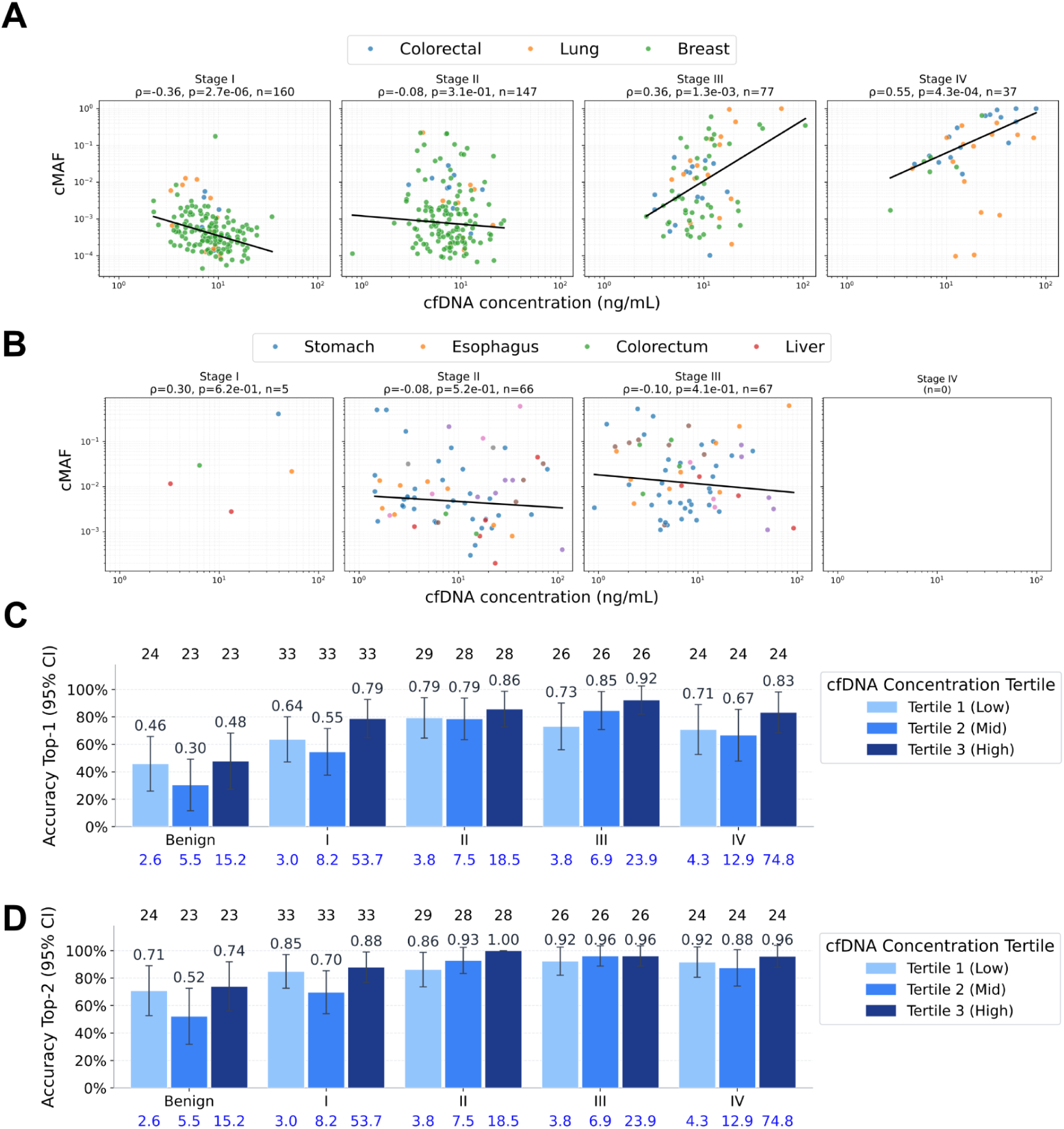
The association between cfDNA concentration, circulating Mutation Allele Frequency (cMAF), and Tissue-of-Origin (TOO) accuracy in published datasets. **(A)** Circulating mutation allele frequency for confirmed somatic variants vs cfDNA concentration from Bredno et al. PLoS One. 2021, stratified by stage. Plots show the Theil-Sen regressor line along with the Spearman coefficient, p value and number of samples. Cancer types in each plot are color coded. **(B)** Same as (A) but for Cohen et al. Science. 2018. **(C)** TOO top-1 accuracy in the independent validation set with 95% CI (bootstrap, n=1000) for cfDNA concentration tertiles within each stage, shown as bar plots. The number of samples in each tertile within each stage is indicated above the plot. The median cfDNA concentration value in ng/mL for each tertile-stage combination is indicated in blue below the bar. **(D)** Same as (C) but top-2 accuracy instead of top-1.

In the Bredno et al. dataset (25), comprising 764 clinically detected and screen-detected cases of breast, lung, and colorectal cancer with staging information available, cMAF data were available for 421 subjects. In these 421 subjects, ctDNA fraction was significantly inversely associated with cfDNA concentration in stage I (ρ=−0.36, p=2.7×10^-6^), non-significantly so in stage II (ρ=−0.08, p=0.3), and significantly positively in later stages (stage III: ρ=0.36, p=0.001; stage IV: ρ=0.55, p=0.0004) (Fig. 6A). Among all 764 cases, cfDNA concentration was not associated with classifier sensitivity after adjusting for cancer type and stage (χ^2^=3.2, df=4, p=0.52) (Supplementary Fig. 7C).

In the Mattox et al. dataset (16), comprising 67 subjects with colorectal, lung, pancreatic, and ovarian cancer, the association between ctDNA fraction and cfDNA concentration was non-significant throughout, negative in stages I–II and near zero in stage III; stage IV had inadequate sample size. Ascertainment data and classifier scores were unavailable for this dataset, precluding analysis of the concentration-sensitivity association (Supplementary Fig. 7A).

The directional consistency across all three datasets is notable: wherever sample sizes permitted evaluation, the correlation between cMAF and cfDNA concentration was negative or non-significant at early stages. Together, these datasets provide no support for the hypothesis that rising cfDNA concentration increases ctDNA fraction in early-stage disease, and the strongest early-stage signal(25) points in the opposite direction. The contrast between Cohen et al. and Bredno et al.—where the concentration-sensitivity association was present in the fully clinically detected cohort but absent in the mixed ascertainment cohort—further suggests that the ascertainment method may modulate this relationship.

We sought additional internal evidence by reasoning that if rising cfDNA concentration were increasing ctDNA fraction, tissue-of-origin (TOO) prediction accuracy—a task that depends directly on the relative abundance of tumor-derived fragments—should also improve with concentration. Fig. 6C and 6D show TOO top-1 and top-2 accuracy, respectively, stratified by cfDNA concentration tertile across stages and the benign cohort. Two patterns are immediately apparent. First, unlike cancer detection sensitivity, TOO accuracy showed no striking monotonic increase across concentration tertiles within most stages. Second, and notably, TOO accuracy in the benign cohort also showed little concentration dependence (top-1: 0.46, 0.30, 0.48 across tertiles; top-2: 0.71, 0.52, 0.74), in contrast to the marked concentration gradient in benign detection sensitivity (Fig. 3C).

Formal testing confirmed the absence of association: after adjusting for cancer type and stage, top-1 accuracy (joint spline test χ^2^=1.58, df=4, p=0.81) and top-2 accuracy (χ^2^=3.1, df=4, p=0.54) showed no concentration dependence in independent validation. Absence of association continued after adjustment for age, gender, BMI, tobacco usage, alcohol usage, plasma storage time, sequencer, DNA input, average coverage and collection time: (χ^2^=2.35, df=4, p=0.67) for top-1 and (χ^2^=3.60, df=4, p=0.46) for top-2 accuracy.

Together, the multiple lines of evidence above suggest that the rise in early-stage sensitivity with increasing cfDNA concentration is not explained by a corresponding increase in ctDNA fraction.

### Small Early-Stage Tumors Show Unexpectedly Elevated cfDNA Concentration

Having established that the concentration-sensitivity association is not explained by ctDNA fraction, we next examined how cfDNA concentration is distributed within the stage categories conventionally used to benchmark MCED performance.

Consistent with previous literature (14–16), cfDNA concentration was elevated in most cancer types relative to controls (Supplementary Fig. 8) and was significantly elevated at every stage relative to controls, including the benign group (Fig. 7A). Unexpectedly, stage I median cfDNA concentration exceeded that of stages II and III (8.2 vs. 7.1 and 6.9 ng/mL respectively), before rising sharply at stage IV (12.9 ng/mL). Stage I also displayed a wider interquartile range than later stages, suggesting substantial within-stage biological heterogeneity. Tumor sizes in our cohort were broadly consistent with AJCC staging expectations (Supplementary Fig. 9B), arguing against systematic understaging as an explanation.

**Figure 7.**
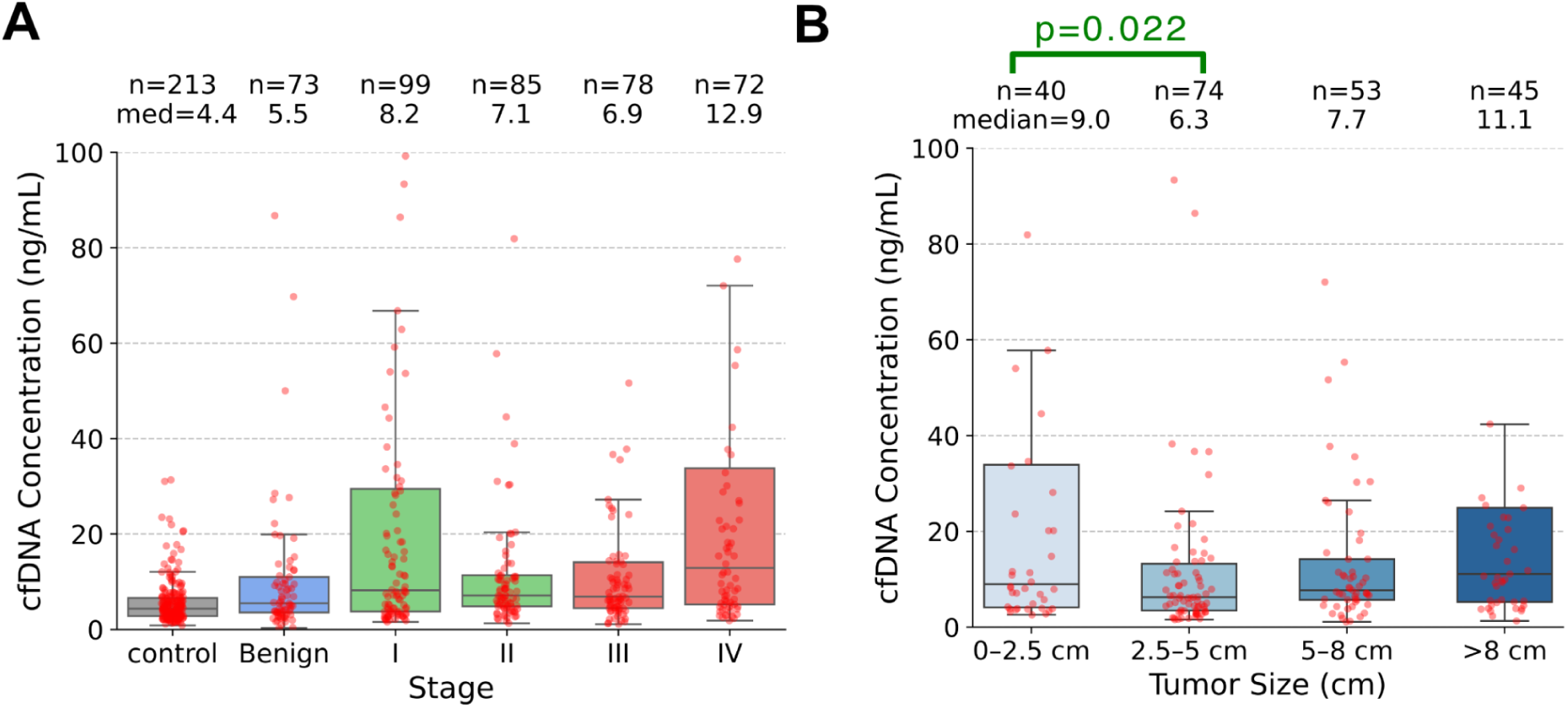
Variation in cfDNA concentration by stage and tumor size in the independent validation and benign cohorts. **(A)** The distribution of concentration stratified by disease stage is shown as box plots, with box medians and sample counts in each group mentioned above the plot. **(B)** The distribution of concentration stratified by tumor size is shown as box plots, with box medians and sample counts in each group mentioned above the plot. Tumor sizes are binned as shown. The two-sided Mann-Whitney p value between the first two bins is also indicated.

Since staging rules vary with cancer type, we examined tumor size as a better proxy for tumor burden. A similar unexpected pattern emerged: tumors smaller than 2.5 cm had higher median cfDNA concentration than those in the 2.5–5 cm range (9.0 vs. 6.3 ng/mL; p=0.022; Fig. 7B), and also exhibited a wider interquartile range.

This pattern is consistent with symptomatic selection bias, whereby smaller tumors presenting symptomatically represent a biologically distinct subset. These findings illustrate that cfDNA concentration, stage, ascertainment method, and tumor biology interact in ways that complicate cross-study benchmarking. Even after standardizing for cancer type, stage, and ascertainment method (Supplementary Results 5), a substantial early-stage sensitivity gap persisted between this study and the CCGA study (2) (stage I: 0.61 vs. 0.25). Further standardization for cfDNA concentration was not possible as concentration data were not reported in CCGA, but the within-stage concentration heterogeneity documented here suggests it may account for part of this difference.

### Symptomatic Benign Lesions and Early Cancers Lie Along a Shared Epigenetic Continuum

The benign cohort had an overall detection rate of 0.33, substantially above the near-zero false positive rate in healthy controls but lower than stage I rates. It also showed a cfDNA concentration-dependent sensitivity gradient comparable in magnitude to that observed in early-stage cancer: detection rate rose from 0.15 in the lowest cfDNA concentration tertile to 0.46 in the highest (Fig. 3C).

The methylation basis for this signal is apparent from the following observations. cfDNA concentrations were significantly elevated relative to controls (Fig. 7A). Ge80 hypermethylation scores in TCGA-derived hypermethylated regions were also significantly elevated in benign lesions relative to healthy controls, at levels intermediate between controls and early-stage cancers (Fig. 4A). Further, these scores grew with cfDNA concentration much like early stage cancers (Fig. 5A). These observations suggest that symptomatic benign lesions share the core epigenetic features that drive cancer classifier signals.

To assess whether the benign and cancer signals are separable, we retrained the classifier with benign samples included as a non-cancer class. Benign detection was reduced to 0%, but overall cancer sensitivity fell from 0.72 to 0.039. The inability to simultaneously maintain high cancer sensitivity and low benign detection suggests that benign lesions and early-stage cancers share substantial methylation biology, creating a challenging trade-off between cancer sensitivity and benign specificity.

## DISCUSSION

In this large multi-institutional Indian case–control study, we demonstrated that total cfDNA concentration is an independent determinant of MCED classifier sensitivity, after rigorous adjustment for cancer type, stage, and demographic and technical covariates. The association was strongest in benign and early stage disease, where cfDNA concentration varied widely within stage and the sensitivity contrast across concentration tertiles was most pronounced, with stage I sensitivity rising monotonically from 0.52 in the lowest tertile to 0.85 in the highest.

Our data extend prior observations that total cfDNA concentration is elevated in malignancy (14–16). This elevation is known to be predominantly leukocyte-derived rather than tumor-derived (16), which would naively predict a dilutive effect on tumor-associated signal. That we observe the opposite—a positive concentration-sensitivity association—implies an increase in the circulating tumor-associated hypermethylated fragment fraction (cTHyF) that drives classifier performance.

Indeed, increased cTHyF was evident in the methylation score data. Higher cfDNA concentration tertiles showed a progressive increase in Ge80 hypermethylation scores specifically within regions identified as differentially hypermethylated in TCGA tumor tissue, while scores for healthy controls remained flat and panel-wide scores declined. Because our classifier relies on precisely these tumor-associated hypermethylated regions to discriminate cancer from non-cancer, their disproportionate enrichment at higher cfDNA concentrations directly explains the concentration-sensitivity association. The declining panel-wide scores, consistent with the known global hypomethylation of cancer tissue (23), further confirm that the region-specific enrichment is biological rather than technical.

Higher ctDNA fraction has been shown to strongly predict detection sensitivity (12,13). Our cohort could not directly adjudicate whether this progressive increase in cTHyF was accompanied by a corresponding increase in ctDNA fraction at early stages. Analysis of three independent published datasets, however, did not support this increase. Across studies, cMAF-estimated ctDNA fraction tended to decrease with increasing cfDNA concentration in early-stage disease. In Bredno et al. (25), this inverse correlation was significant at stage I (ρ = −0.36, p = 2.7 × 10^-6^) and weaker but directionally consistent at stage II, while Cohen et al. and Mattox et al. (11,16) reported similar negative correlations that did not reach statistical significance. Further support came from the observation that the association between cfDNA concentration and assay performance appeared specific to cancer detection rather than tissue-of-origin (TOO) prediction. Detection sensitivity is based on tumor-derived hypermethylation signals, while TOO prediction may depend more on lineage-associated signals (26). Therefore, if cfDNA concentration increases were accompanied by substantially rising ctDNA fraction, lineage-specific signals and TOO accuracy would be expected to rise in parallel; yet TOO accuracy showed no clear concentration dependence, providing additional evidence against a rising ctDNA fraction at early stages.

To explain the rise in cTHyF without a rise in ctDNA fraction at early stages, we propose that two distinct phenomena are at play. The first is a systemic axis (16) where increasing cfDNA concentration as a result of generalized tissue and hematopoietic cell turnover and DNA clearance is associated with an increasing intensity of epigenetic dysregulation in the tumor, thus enriching cTHyF independently of ctDNA fraction and accentuating detection. The second is a local axis (27,28) where increasing cfDNA concentration simply dilutes the ctDNA fraction that is governed by apoptosis, necrosis, secretion, and vascular dissemination pathways (29), thus attenuating detection. These axes likely operate in varying combinations across cohorts: screen-detected and mixed-ascertainment cohorts enriched for biologically quiescent tumors (e.g., Bredno et al.) may lean toward the local-axis-dominant dilutive pattern, explaining the absence of a concentration-sensitivity association in Bredno et al. Symptomatic or clinically detected cohorts enriched for biologically active tumors (e.g., this cohort and Cohen et al. (11)) may lean toward the systemic-axis-dominant dysregulation-driven pattern, consistent with the significant positive concentration-sensitivity associations observed in both.

Beyond early-stage tumors, a parallel pattern was evident for benign lesions, suggesting that systemic axis engagement is not exclusive to malignancy. cfDNA concentrations, and Ge80 hypermethylation scores in TCGA-derived regions, were significantly elevated in an independent symptomatic benign cohort at levels intermediate between healthy controls and early-stage cancers. These scores also broadly increased with cfDNA concentration. Classifier positivity rates reflected this gradient, rising from 0.15 in the lowest concentration tertile to 0.46 in the highest, suggesting that the same enrichment phenomenon observed in early-stage cancer may also be active in symptomatic benign conditions. These findings have direct clinical implications. Systematic evaluation of symptomatic benign disease has been largely absent from published MCED studies, yet our data, together with prior work in ovarian cancer (30), suggest that distinguishing benign from malignant conditions at high specificity represents a substantive challenge for methylation-based MCED assays in symptomatic populations, and that downstream diagnostic workup will likely be necessary in such settings.

Beyond their biological implications, our findings have clinical implications in cross-study benchmarking. Our cohort demonstrated stage I sensitivity of 0.72 at 0.986 specificity, comparable to the performance reported in several studies (3,7,8) and higher than reported in CCGA (2). The differences in sensitivity with CCGA persisted even after standardization for cancer type, stage, and ascertainment, pointing to residual sources of variability that remain incompletely accounted for. Concentration-standardized comparisons may help explain some of these differences; however, such analyses are currently precluded by the absence of sample-level cfDNA concentration and ascertainment data in CCGA. Nevertheless, our results support the routine reporting of cfDNA concentration data in MCED studies.

Our study has the following limitations. Cases were largely symptomatic; early-stage symptomatic tumors, in particular, are likely enriched for biological activity which may have amplified the concentration-sensitivity association relative to what would be observed in screen-detected cohorts where small tumors are more likely to be biologically quiescent. The study did not address standardization of concentration measurements across varying extraction and quantification methods. The relationship between tumor epigenetic dysregulation and cfDNA concentration has not been causally established in this study. The association between ctDNA fraction and cfDNA concentration was not directly assessed in this cohort, and inferences made were indirect. Sample sizes for several individual cancer types were limited, restricting high-confidence analyses stratified simultaneously by cancer type, stage, and concentration. Finally, although robustness analyses demonstrated stability across sites, time points, and sequencing batches, broader external validation would strengthen these conclusions.

Future research should prioritise: prospective validation of the concentration-sensitivity relationship in asymptomatic screening cohorts, where the dilutive local axis may dominate; direct measurement of ctDNA fraction alongside total cfDNA concentration to mechanistically dissect the relative contributions of the systemic and local axes; development of concentration-reweighting approaches to estimate sensitivity attenuation when translating from symptomatic to screening settings; longitudinal follow-up of benign detections to determine their clinical trajectory and inform the design of downstream diagnostic algorithms; and external validation of these findings in geographically and demographically distinct populations.

## CONCLUSIONS

This study establishes total cfDNA concentration as an independent determinant of MCED classifier sensitivity in a large, multi-institutional Indian cohort, grounded in the enrichment of tumor-associated hypermethylated fragments with rising cfDNA concentration. The dissociation between concentration-dependent gains in detection sensitivity and the stability of tissue-of-origin accuracy, together with evidence from independent external datasets, suggests that this enrichment is not driven by a concomitant rise in ctDNA fraction. We propose a two-axis model to explain these findings—a systemic axis where greater cfDNA concentration reflects higher epigenetic dysregulation in the tumor independently of ctDNA fraction and thus accentuates detection, and a local axis where greater cfDNA concentration dilutes ctDNA fraction and attenuates detection. Symptomatic and screen-detected cohorts are likely to engage these axes in different proportions, with implications for how concentration-sensitivity relationships and sensitivity estimates should be interpreted and compared across studies. The parallel engagement of this mechanism in symptomatic benign disease further underscores the biological continuity between benign and malignant epigenetic dysregulation, and highlights the diagnostic challenge of distinguishing these conditions at high specificity in symptomatic populations. Taken together, these findings suggest that cfDNA concentration, alongside cancer type, stage, ctDNA fraction, and ascertainment method, should be considered an explicit variable in the design, analysis, and benchmarking of methylation-based MCED assays.

## Supporting information

Supplementary methods and results

Sample-level details for both the main cohort and the benign cohort

TCGA beta values for 4384 classifier regions and for the entire TCGA panel

Classifier performance sensitivity/specificity/AUC/CIs (Main + Auxiliary Splits)

Confounder analysis

TOO results

Sample-wise GE80/LT20 across for TCGA-derived regions

Tumor fraction (cMAF) vs cfDNA concentration analysis

Benchmarking Across Studies

## Data Availability

The data generated in this study contain potentially identifiable genomic and clinical information and are therefore not publicly available. De-identified data supporting the findings of this study, including processed feature matrices and summary-level results, are available from the corresponding authors upon reasonable request and subject to institutional review, data use agreements, and applicable ethical and regulatory approvals. Detailed derived data is included in the supplementary tables, and code is included at our github repository at this link to enable reproducibility: https://github.com/ShashankStrand/Strand_MCED_EMERGE.

## ABBREVIATIONS

cfDNA: Cell-free DNA
ctDNA: Circulating tumor DNA
cMAF: Circulating mutant allele frequency
cTHyF: Circulating tumor-associated hypermethylated fragment fraction
MCED: Multi-cancer early detection
TOO: Tissue of origin
Ge80: The fraction of hypermethylated fragments
Lt20: The fraction of hypomethylated fragments
AUC: Area under the curve
ROC: Receiver operating characteristic

## DECLARATIONS

### 1. Ethics approval and consent to participate

In the EMERGE study (Epigenetic Markers for Early and Robust Cancer Detection in a General Population), the names of the Ethics Committees that approved the study and their respective registration numbers appear below. In total, 22 IEC approvals were obtained covering 43 recruitment sites. Informed consent was obtained from all individual participants included in the study.

1. Ethiclin Pvt Ltd IEC: ECR/350/Indt/KA/2021
2. HCG-BBHIO Ethics Committee: ECR/994/INST/KA/2017/RR-21
3. BGS Global Hospitals IEC: ECR/128/Inst/KA/2013/RR-19
4. Prakriya Hospitals IEC: ECR/1412/Inst/KA/2020
5. Shreyas Hospital IEC: ECR/962/Inst/MH/2017/RR-20
6. Shanmuga Medical Research Foundation Trust IEC: ECR/1046/Inst/TN/2018/RR-21
7. Erode Cancer Centre IEC: ECR/319/INST/TN/2013/RR-19
8. CARE Multispeciality Hospitals IEC: ECR/94/Inst/AP/2013/RR-21
9. Shrey Hospital IEC: ECR/1302/Inst/GJ/2019
10. HP Poddar Memorial Clinic and Nursing Home IEC: ECR/1555/Inst/WB/2021
11. Manavata Research Center IEC: ECR/500/Inst/MH/2013/RR-20
12. Amravati Ethics Committee, SSCHACF: ECR/432/Inst/MH/2013/RR-19
13. Galaxy Care Multispeciality Hospital IEC: ECR/1379/Inst/MH/2020
14. Aurangabad Health Care and Research LLP IEC: ECR/325/Indt/MH/2020
15. Telerad RxDx Healthcare Pvt Ltd IEC: ECR/1494/Inst/KA/2021
16. HCG-Central Ethics Committee: ECR/386/Inst/KA/2013/RR-19
17. Sangini Hospital IEC: ECR/147/Inst/GJ/2013/RR-19
18. Krishna Institute of Medical Sciences "Deemed to be University" IEC: ECR/307/Inst/MH/2013/RR
19. Shri Siddhivinayak Ganapati Cancer Hospital IEC: ECR/588/Inst/MH/2014/RR-21
20. Chittaranjan National Cancer Institute IEC: EC/NEW/INST/2020/946
21. Saveetha Medical College and Hospital IEC: ECR/724/Inst/TN/2015/RR-24
22. Central IEC: ECR/390/Indt/MH/2024

### 2. Consent for publication

Not applicable. At enrolment all samples were de-identified, participants signed informed consent. The manuscript does not contain any individual person’s data in any form (including individual details, images, or videos).

### 6. Authors’ contributions

S.B., A.C., V.V., R.H.: conceived the study, supervised the project, performed troubleshooting, interpreted results, accessed and verified the data, and wrote the manuscript with input from all authors; A.C., A.G., A.R., S.U., A.K., L.B., P.G., A.S.: obtained regulatory approvals, recruited subjects, and managed samples and data; D.V., P.R.T., A.N., A.B., S.P.: validated and executed assay protocols; P.H., N.R., T.S., S.K., B.P., L.N., K.S., K.Se., S.R., Y.K.S.: performed data analysis and software development; S.Y., B.G., U.B.: conducted literature review; C.R.C., S.L., V.C.: provided conceptual guidance and expert oversight; All authors had full access to all the data in the study, reviewed and approved the final manuscript, and had final responsibility for the decision to submit for publication.

## 7. Acknowledgements

The results shown here are in whole or part based upon data generated by the TCGA Research Network: https://www.cancer.gov/tcga

AI-based language assistance tools were used for language editing and manuscript polishing. We thank Varun Aggarwala and Ankita Bansal for critical reviews of the manuscript and suggestions for improvement. We thank V.K. Chaithanya Ponnaluri and Ashwani Kamal for insights into standardization of the EM-seq protocol. We thank Niranjan Nagarjan for a critical reading of the manuscript and his many suggestions. We also thank the clinicians and institutions involved in participant recruitment and sample collection for their contributions. Finally, we thank our colleagues at Strand Life Sciences Pvt. Ltd. for numerous helpful discussions and support throughout the project.

## 8. Authors’ information (optional)

V.C. and R.H. are ex-faculty members of the Indian Institute of Science. V.C is a Commissioner with the Lancet Citizens Commission for reimagining India’s health systems. R.H is a fellow of the Indian Academy of Sciences and the Indian National Academy of Engineering. R.H is the author of *Genomic Quirks*, published by the Indian Institute of Science press. C.R.C is an ex-faculty member of Boston University, Columbia University, and University of California, Berkeley. C.R.C co-authored *Biophysical Chemistry* with Paul Schimmel and *Genomics: The Science and Technology Behind the Human Genome Project* with Cassandra Smith. A.C. is an adjunct faculty at Department of Oncology, Johns Hopkins University, Baltimore, MD. S.L is an MD, and Director Medical & Precision Oncology and Director Oncology Research, Sir HN Reliance Foundation Hospital & Research Center.

## REFERENCES

1. Liu MC, Oxnard GR, Klein EA, Swanton C, Seiden MV, CCGA Consortium. Sensitive and specific multi-cancer detection and localization using methylation signatures in cell-free DNA. Ann Oncol. 2020 Jun;31(6):745–59.

2. Klein EA, Richards D, Cohn A, Tummala M, Lapham R, Cosgrove D, et al. Clinical validation of a targeted methylation-based multi-cancer early detection test using an independent validation set. Ann Oncol. 2021 Sep;32(9):1167–77.

3. Bao H, Yang S, Chen X, Dong G, Mao Y, Wu S, et al. Early detection of multiple cancer types using multidimensional cell-free DNA fragmentomics. Nat Med. 2025 Aug;31(8):2737–45.

4. Chung DC, Gray DM 2nd, Singh H, Issaka RB, Raymond VM, Eagle C, et al. A Cell-free DNA Blood-Based Test for Colorectal Cancer Screening. N Engl J Med. 2024 Mar 14;390(11):973–83.

5. Mathios D, Niknafs N, Annapragada AV, Bobeff EJ, Chiao EJ, Boyapati K, et al. Detection of Brain Cancer Using Genome-wide Cell-free DNA Fragmentomes. Cancer Discov. 2025 Aug 4;15(8):1593–608.

6. Zhang X, Wang Z, Tang W, Wang X, Liu R, Bao H, et al. Ultrasensitive and affordable assay for early detection of primary liver cancer using plasma cell-free DNA fragmentomics. Hepatology. 2022 Aug;76(2):317–29.

7. Nguyen VTC, Nguyen TH, Doan NNT, Pham TMQ, Nguyen GTH, Nguyen TD, et al. Multimodal analysis of methylomics and fragmentomics in plasma cell-free DNA for multi-cancer early detection and localization. Elife [Internet]. 2023 Oct 11;12. Available from: 10.7554/eLife.89083

8. Li S, Geng S, Chen Y, Ren Q, Luan Y, Liang W, et al. Clinical Validation of a Noninvasive Multi-Omics Method for Multicancer Early Detection in Retrospective and Prospective Cohorts. J Mol Diagn. 2025 Jul;27(7):657–70.

9. Jiang P, Sun K, Peng W, Cheng SH, Ni M, Yeung PC, et al. Plasma DNA End-Motif Profiling as a Fragmentomic Marker in Cancer, Pregnancy, and Transplantation. Cancer Discov. 2020 May;10(5):664–73.

10. Cristiano S, Leal A, Phallen J, Fiksel J, Adleff V, Bruhm DC, et al. Genome-wide cell-free DNA fragmentation in patients with cancer. Nature. 2019 Jun;570(7761):385–9.

11. Cohen JD, Li L, Wang Y, Thoburn C, Afsari B, Danilova L, et al. Detection and localization of surgically resectable cancers with a multi-analyte blood test. Science. 2018 Feb 23;359(6378):926–30.

12. Bredno J, Venn O, Chen X, Freese P, Ofman JJ. Circulating Tumor DNA Allele Fraction: A Candidate Biological Signal for Multicancer Early Detection Tests to Assess the Clinical Significance of Cancers. Am J Pathol. 2022 Oct;192(10):1368–78.

13. Jamshidi A, Liu MC, Klein EA, Venn O, Hubbell E, Beausang JF, et al. Evaluation of cell-free DNA approaches for multi-cancer early detection. Cancer Cell. 2022 Dec 12;40(12):1537–49.e12.

14. Huang RSP, Xiao J, Pavlick DC, Guo C, Yang L, Jin DX, et al. Circulating Cell-Free DNA Yield and Circulating-Tumor DNA Quantity from Liquid Biopsies of 12 139 Cancer Patients. Clin Chem. 2021 Nov 1;67(11):1554–66.

15. Leon SA, Shapiro B, Sklaroff DM, Yaros MJ. Free DNA in the serum of cancer patients and the effect of therapy. Cancer Res. 1977 Mar;37(3):646–50.

16. Mattox AK, Douville C, Wang Y, Popoli M, Ptak J, Silliman N, et al. The Origin of Highly Elevated Cell-Free DNA in Healthy Individuals and Patients with Pancreatic, Colorectal, Lung, or Ovarian Cancer. Cancer Discov. 2023 Oct 5;13(10):2166–79.

17. Erger F, Nörling D, Borchert D, Leenen E, Habbig S, Wiesener MS, et al. cfNOMe - A single assay for comprehensive epigenetic analyses of cell-free DNA. Genome Med. 2020 Jun 24;12(1):54.

18. Bie F, Wang Z, Li Y, Guo W, Hong Y, Han T, et al. Multimodal analysis of cell-free DNA whole-methylome sequencing for cancer detection and localization. Nat Commun. 2023 Sep 27;14(1):6042.

19. Vaisvila R, Ponnaluri VKC, Sun Z, Langhorst BW, Saleh L, Guan S, et al. Enzymatic methyl sequencing detects DNA methylation at single-base resolution from picograms of DNA. Genome Res. 2021 Jul;31(7):1280–9.

20. Nuttall B, Karl DL, Burke K, Callahan M, Mendler K, Cingolani P, et al. Comprehensive comparison of enzymatic and bisulfite DNA methylation analysis in clinically relevant samples. Clin Epigenetics. 2025 Oct 3;17(1):156.

21. Amin MB, Greene FL, Edge SB, Compton CC, Gershenwald JE, Brookland RK, et al. The Eighth Edition AJCC Cancer Staging Manual: Continuing to build a bridge from a population-based to a more “personalized” approach to cancer staging. CA Cancer J Clin. 2017 Mar;67(2):93–9.

22. Edge SB, Compton CC. The American Joint Committee on Cancer: the 7th edition of the AJCC cancer staging manual and the future of TNM. Ann Surg Oncol. 2010 Jun;17(6):1471–4.

23. Stackpole ML, Zeng W, Li S, Liu CC, Zhou Y, He S, et al. Cost-effective methylome sequencing of cell-free DNA for accurately detecting and locating cancer. Nat Commun. 2022 Sep 29;13(1):5566.

24. Aerden M, Jatsenko T, Leroy K, De Ridder K, Nootens A, Piatti V, et al. Longitudinal cell-free DNA methylome and fragmentome profiles in health uncover signatures of cell type and demographic origin. Genome Med [Internet]. 2026 Apr 9;18(1). Available from: 10.1186/s13073-026-01618-w

25. Bredno J, Lipson J, Venn O, Aravanis AM, Jamshidi A. Clinical correlates of circulating cell-free DNA tumor fraction. PLOS ONE. 2021 Aug 25;16(8):e0256436.

26. Moss J, Magenheim J, Neiman D, Zemmour H, Loyfer N, Korach A, et al. Comprehensive human cell-type methylation atlas reveals origins of circulating cell-free DNA in health and disease. Nature Communications. 2018 Nov 29;9(1):5068.

27. Ozyerli-Goknar E, Bagci-Onder T. Epigenetic Deregulation of Apoptosis in Cancers. Cancers (Basel) [Internet]. 2021 Jun 27;13(13). Available from: 10.3390/cancers13133210

28. Zhang L, Liang Y, Li S, Zeng F, Meng Y, Chen Z, et al. The interplay of circulating tumor DNA and chromatin modification, therapeutic resistance, and metastasis. Mol Cancer. 2019 Mar 9;18(1):36.

29. Avanzini S, Kurtz DM, Chabon JJ, Moding EJ, Hori SS, Gambhir SS, et al. A mathematical model of ctDNA shedding predicts tumor detection size. Sci Adv [Internet]. 2020 Dec;6(50). Available from: 10.1126/sciadv.abc4308

30. Medina JE, Annapragada AV, Lof P, Short S, Bartolomucci AL, Mathios D, et al. Early Detection of Ovarian Cancer Using Cell-Free DNA Fragmentomes and Protein Biomarkers. Cancer Discov. 2025 Jan 13;15(1):105–18.

