## Supplementary methods and results for "cfDNA concentration as an independent determinant of multi-cancer early detection sensitivity: evidence from a large Indian case-control cohort"

### SUPPLEMENTARY RESULTS

#### 1. Classifier Characteristics

ROC analysis demonstrated discrimination between cancer cases and controls in cross-validation (AUC: 0.967; 95% CI 0.955–0.977) and in independent validation (AUC: 0.960; 95% CI 0.947–0.973).

At the pre-specified 99% training specificity threshold, specificity in the validation cohort was 0.986 (95% CI 0.968–1) while overall sensitivity was comparable between training and validation cohorts (0.69 vs 0.72, respectively). Sensitivity for stage I was 0.72 (95% CI 0.63–0.80) in the validation cohort and remained comparable across stages II and III (0.67 and 0.72, respectively) before increasing in stage IV to 0.83.

Cancer type-specific analyses in the validation cohort demonstrated high sensitivity in liver, lung and esophageal cancers (Supplementary Fig. 3) consistent with prior reports (3). In the symptomatic benign cohort, the detection rate was 0.33 (95% CI 0.24–0.44).

The fragment-level rescue step that selectively removed poorly converted fragments from all samples removed ~13% of fragments without bias in both cases and controls and restored >99% non-CpG cytosine conversion (Supplementary Results 2).

Classifier performance remained robust to major sources of technical, geographic, temporal, and demographic variability that could potentially confound concentration-sensitivity association analyses (Supplementary Results 3).

Tissue-of-origin (TOO) top-1 prediction accuracy after grouping cancers into nine anatomically related categories to ensure adequate sample sizes was 0.74 (95% CI 0.70–0.77) in cross-validation and 0.76 (95% CI 0.71–0.80) in independent validation (Supplementary Fig. 3), increasing to 0.87 (95% CI 0.84–0.89) and 0.90 (95% CI 0.86–0.93), respectively, for top-2 predictions. Restricting analysis to cancer detection–positive samples produced a ~0.02 improvement in top-1 accuracy and minimal change in top-2 accuracy.

Details are in Supplementary Tables 3 and 5.

#### 2. Fragment-level Rescue Retains Specificity without Reruns

A technical challenge with enzymatic conversion is inadequate conversion of non-CpG cytosines (24). 20.4% of cases and 22.4% of controls had less than 99% non-CpG cytosine conversion in our cohort. We implemented a fragment-level rescue step that selectively removed poorly converted fragments from all samples, rather than discard these failed samples. This reduced usable fragments by a median of 13% in both groups without evidence of differential bias (p=0.97), while restoring the 99% non-CpG cytosine conversion threshold to all samples. Without this rescue step, proceeding with the failed samples inflated classifier scores in controls and reduced specificity in the independent validation set by 6% with only a marginal sensitivity gain. These findings indicate that fragment-level rescue is an effective strategy for maintaining assay specificity without reruns in enzymatic methyl-sequencing workflows (Supplementary Fig. 10).

##### 3. Adversarial Splits & Adjustment for Covariates Suggest Classifier Robustness

To assess classifier robustness, we considered two related but distinct questions: (1) deployment robustness: whether sufficient covariate heterogeneity was present to evaluate stability under real-world variation, and (2) confounding: whether covariates could induce confounding methylation patterns associated with an apparent cancer signal.

###### Deployment robustness

Performance was consistent across training-validation splits stratified in turn by sequencing batch, collection time, and recruitment site, with sensitivity ranging from 0.66–0.73 and specificity from 0.97–1.00 (Supplementary Fig. 11). Site-wise splits were constructed such that each site contributed exclusively to either training or validation, preventing geographic leakage across splits. Similarly sequencing batch splits were created so each reagent batch contributed only to training or validation, preventing technical leakage across splits. This suggests that the classifier is robust to changes in reagents, sites and temporal factors expected in real life.

We note that all samples were processed through a single central laboratory under a tightly controlled common protocol regardless of case/control status or recruitment site; laboratory personnel, instruments, and core reagents were therefore shared across splits. This shared infrastructure is unlikely to introduce differential leakage because processing protocols were identical across groups, but it does mean these splits test robustness to temporal, geographic, and batch variability within a common infrastructure rather than true external generalizability. This is an inherent limitation of single-centre processing studies, and we regard independent external validation as an important direction for future work. Nonetheless, the current splits provide the appropriate validity evidence for the intended deployment model, in which the test will be run from a single central laboratory serving geographically diverse collection sites, precisely the configuration these splits were designed to stress-test.

###### Site imbalance and geographic confounding

Cases were recruited from 40 sites and controls from 3 sites, raising the concern of geographic constraint bias. However, several lines of evidence argue against such a bias.

First, the three control sites were geographically diverse, one each in the west, south, and east of India (Fig. 1A), mirroring the distribution of sites where cases were recruited, and recruitment spanned several months at each site, introducing further heterogeneity.

Second, to assess whether the site imbalance introduced a systematic methylation artifact, we leveraged bidirectional concordance with TCGA, an entirely independent dataset with different sites, collection protocols, and processing infrastructure. Ge80 hypermethylation scores in

TCGA-derived cancer-associated regions were elevated in our cases (Fig. 4). Conversely, regions identified as hypermethylated in our cohort showed concordant differential methylation in TCGA (Fig. 2A). Together, these reciprocal findings across independent datasets with distinct collection and processing frameworks argue against site-specific artifact as the primary explanation for classifier performance.

Third, we assess the differential contribution of the 3 control recruitment sites. Ge80 hypermethylation scores in TCGA-derived cancer-associated regions were elevated in our cases relative to each of the 3 sites (Supplementary Fig. 12). Classifier score distributions varied modestly between control sites but were concentrated well below the detection cutoff for each of the 3 sites; only 0%, 1.5%, and 2.2% of samples, respectively, achieved scores above the cutoff.

**Covariate confounding**

Cases and controls differed on some demographic and technical variables (Table 1), including sex (47.1% vs 57.3% male), tobacco usage (22.3% vs 69.8%), plasma storage time (median 218 vs. 71 days) and sequencer (97.4% vs. 69.1% NovaSeq X+), making adjustment for these variables particularly important. Progressive covariate adjustment showed that the classifier-cancer association was robust across all models (Supplementary Table 4). The unadjusted odds ratio was 399.7 (95% CI 234.8–680.4); sequential adjustment for age, sex, BMI, alcohol use, plasma storage time, and sequencer yielded OR 360.4 (95% CI 195.9–662.8), and further adjustment for tobacco use yielded OR 436.2 (95% CI 206.8–920.3), with overlapping confidence intervals across all models. The E-value for the fully adjusted OR was ~871, indicating substantial robustness to potential unmeasured confounding.

To assess sensitivity to covariate imbalance more broadly, inverse probability weighting (IPW) was applied across the seven binarized covariates. Weighted ORs ranged from 417.1 (flow cell) to 625.6 (tobacco), all remaining highly statistically significant. IPW achieved balance (standardized mean differences <0.12) for most covariates; residual imbalance after sequencer weighting (plasma storage time SMD 0.80) and alcohol weighting (tobacco usage SMD 0.37, gender SMD 0.4) reflects collinearity between these variables, which were independently adjusted for in the regression model. Together, these analyses suggest that the classifier-cancer association is unlikely to be materially explained by imbalance in the evaluated covariates.

**Further analysis of Tobacco as a potential confounder**

Since controls were substantially enriched for tobacco users relative to cases (69.8% vs. 22.3%), we performed additional analyses stratified by tobacco use status. Strong associations of classifier score with case-control status were observed in both tobacco users (OR 990.2, 95% CI 285.6–3433.5) and non-users (OR 267.6, 95% CI 95.5–750.0), with no evidence of interaction between score and tobacco usage status (p=0.23).

False positive rates showed overlapping confidence intervals between tobacco strata across all training specificity thresholds with no systematic directional difference (Supplementary Fig. 13A).

True positive rates were numerically higher in tobacco users (Supplementary Fig. 13B) consistently across thresholds. However, this separation was not evident within most individual cancer types, including lung and esophageal, the cancers most strongly associated with tobacco; these showed little or no difference or had strongly overlapping confidence intervals. The aggregate true positive rate difference therefore was largely attributable to enrichment of high-sensitivity cancer types, particularly lung and esophageal cancers, within the tobacco-user subgroup (lung and esophageal tobacco user fractions of 47% and 42% respectively). Taken together, these analyses suggest that tobacco enrichment among controls does not materially confound classifier performance.

**4. Classifier Regions Enrich for Pan-Cancer Hypermethylation Patterns**

The Twist Methylome Panel comprises 551,803 targeted regions of which our classifier used 4,384 regions. Compared with the broader 150k background subset of the 550K regions with at least 5 CpGs, the 4,384 classifier regions showed distinct genomic characteristics. Classifier regions were substantially longer, with 51% exceeding 500 bp in length versus 26.3% in the 150k background set. They were also more enriched for promoter regions (55% versus 46.9%) and exhibited higher CpG density, averaging 6.3 CpGs per 100 bases compared with 4.9 CpGs per 100 bases in the 150k set.

Functional enrichment analysis of promoter genes associated with classifier regions (Supplementary Fig. 14) using Reactome 2024 pathways identified 13 enriched terms that were absent across all 10 matched background iterations, with odds-ratio enrichments reaching approximately 13-fold. These pathways were predominantly related to developmental regulation, neuronal lineage specification, and transcriptional silencing programs.

Epigenomics Roadmap analysis of promoter genes associated with classifier regions identified 14 enriched terms dominated by H3K27me3- and H3K9me3-associated chromatin states across epithelial and immune cell lineages. These findings are consistent with prior models in which Polycomb-associated H3K27me3-mediated repression may predispose genomic regions to more stable H3K9me3- and DNA methylation-associated silencing during tumorigenesis.

Analysis of transcription factor motifs and tissue-specific enhancers did not result in significant enrichment. These findings suggest that classifier regions do not primarily reflect tissue-restricted regulatory programs, but instead capture broadly shared cancer-associated hypermethylation events during tumorigenesis.

Finally, since differences in chromatin accessibility across genomic regions manifest as differences in fragmentation patterns in cfDNA resulting in sequencing depth variation, we assessed the discriminative ability of our Ge80 hypermethylation score versus that of normalized sequencing depth. Both the 4,384 classifier regions as well as matched random background regions showed small effect sizes for differences in normalized sequencing depth between cases and controls (Cohen's d depth median = −0.122, and median = −0.076, respectively). In contrast, Ge80 scores showed stronger positive effect sizes in the classifier regions (Cohen's d Ge80 median = +0.503) than in matched random background regions (median = +0.320, respectively). While fragmentation and methylation are likely coupled consequences of the same underlying chromatin state, these findings support tumour-associated hypermethylation constituting the dominant and biologically selective discriminative signal captured (Supplementary Fig. 15).

##### 5. Stage, Cancer Type & Ascertainment Do Not Fully Explain Between-study Sensitivity Differences

To assess whether conventional benchmarking variables fully explain cross-study performance variation, we compared the sensitivity of our assay against the Circulating Cell-Free Genome Atlas (CCGA) study (2).

While both studies were case-control studies, cancer type and stage distributions varied between the studies. In addition, the mode of ascertainment also varied, with 73% of CCGA cases and almost all cases in this study being diagnosed upon clinical presentation, and 27% of cases in CCGA diagnosed via screening. Neither study tracked incidental detection rates; however, population estimates suggest these contribute minimally to overall ascertainment composition (Koo et al. *BMJ Open* 2019).

To standardize ascertainment, we restricted analysis to nine non-screenable cancer types diagnosed upon clinical presentation (and not via screening) in both cohorts (Supplementary Table 8). A substantial early-stage sensitivity gap persisted after standardization of cancer type and stage distributions: our Stage I sensitivity was substantially higher than CCGA (0.61 vs. 0.25), while Stage II sensitivity differences were less pronounced (0.65 vs. 0.53) (Supplementary Fig. 9A).

Tumor sizes in our cohort were broadly consistent with AJCC staging expectations (Supplementary Fig. 9B), arguing against systematic understaging as the primary explanation for the observed early-stage detection discrepancy. The substantial heterogeneity in cfDNA concentration observed within our stage I cohort motivates consideration of cfDNA concentration as a potential contributor to this discrepancy. However, cfDNA concentration distributions were not reported in CCGA, precluding a direct concentration-standardized comparison.

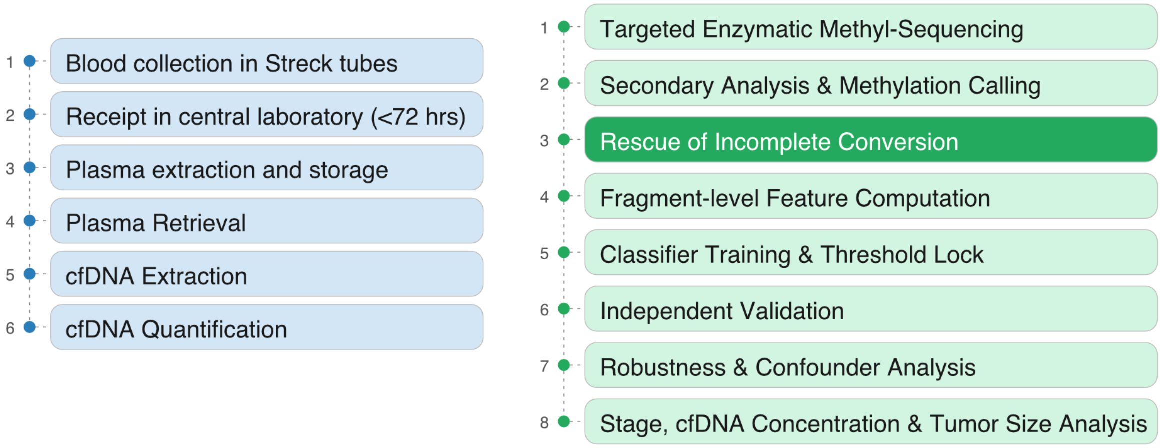


**Supplementary Figure 1. Pre-analytical & Analytical Workflow:** The key sample collection and transport steps, and the main steps of our sequencing and analysis workflow.

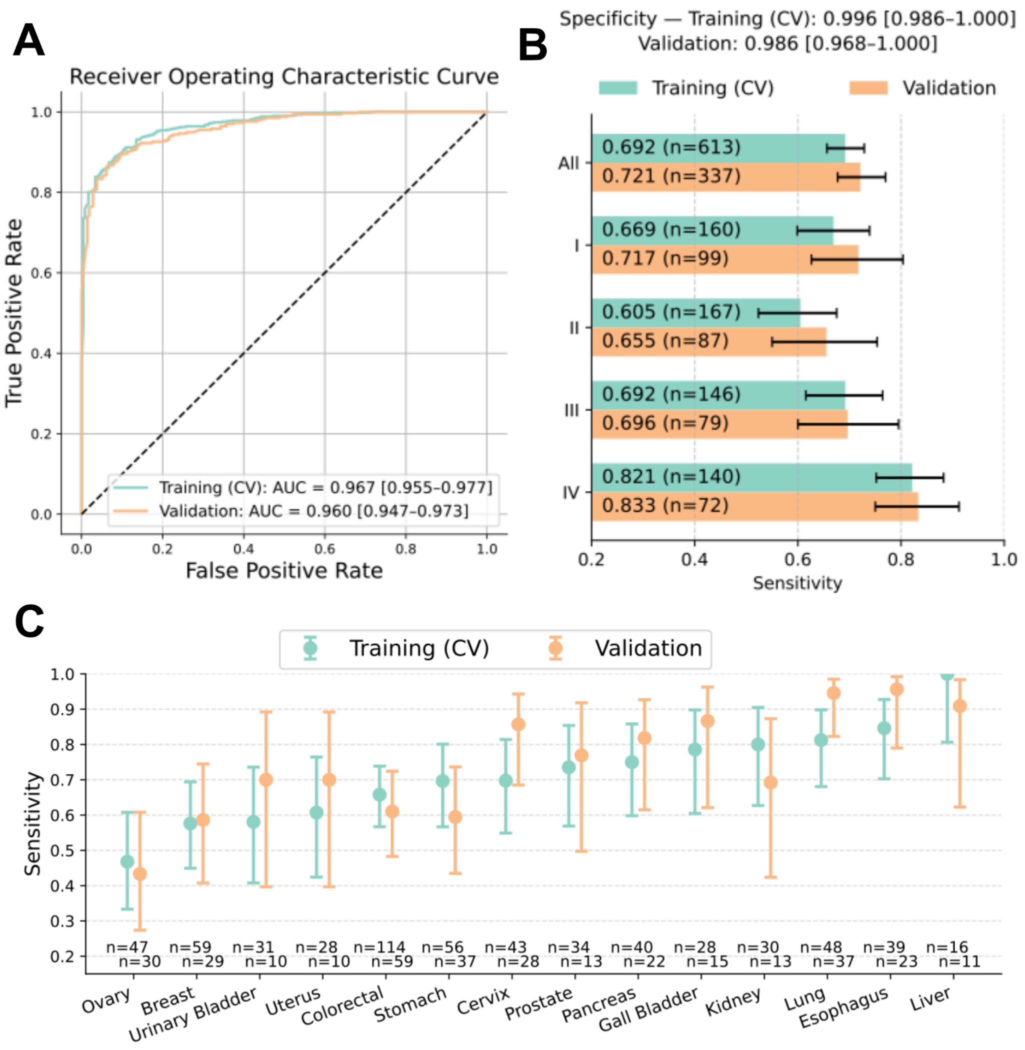


**Supplementary Figure 2. Classifier performance in our main cohort. (A)** True positive rate versus false positive rate curves for cross-validation (on the training set) and for independent validation are shown, along with AUC values and 95% CIs (derived by bootstrap, n=1000). **(B)** Cross-validation and independent validation sensitivities are indicated as bar plots by stage, at the pre-specified 99% training specificity cutoff. The number of samples and 95% CIs are also indicated (derived by bootstrap, n=1000). **(C)** At the pre-specified 99% training specificity cutoff**,** sensitivity by cancer type in cross-validation and in independent validation is indicated as dot plots for the 14 cancer types in our main cohort; error bars show 95% CIs (derived by bootstrap, n=1000). The number of samples in each case is also indicated.

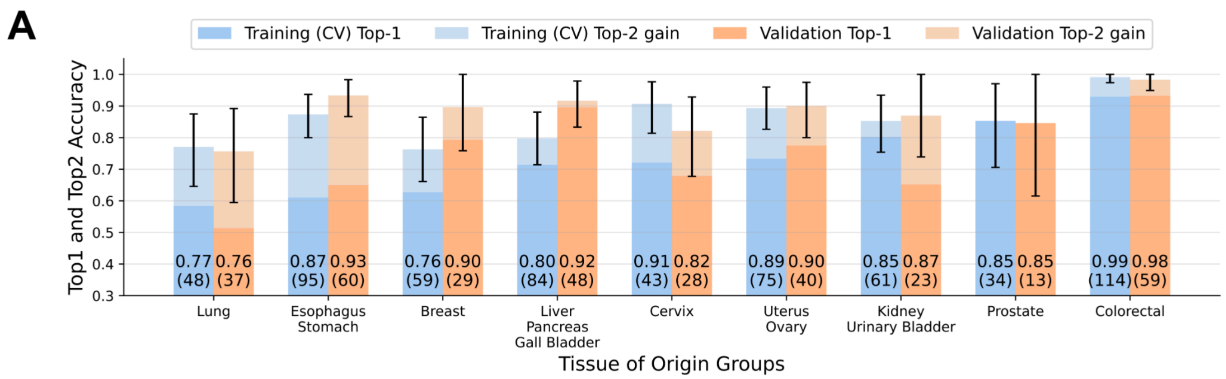


**Supplementary Figure 3. Tissue-of-Origin (TOO) prediction. (A)** TOO performance in cross-validation and in independent validation as bar plots, with both top-1 and top-2 performance shown in different shades. Top-2 performance is shown as a gain over top-1. 95% CIs (bootstrap, n=1000) are also shown. For each bar, the bar value is indicated in text and the associated sample count is mentioned in round brackets.

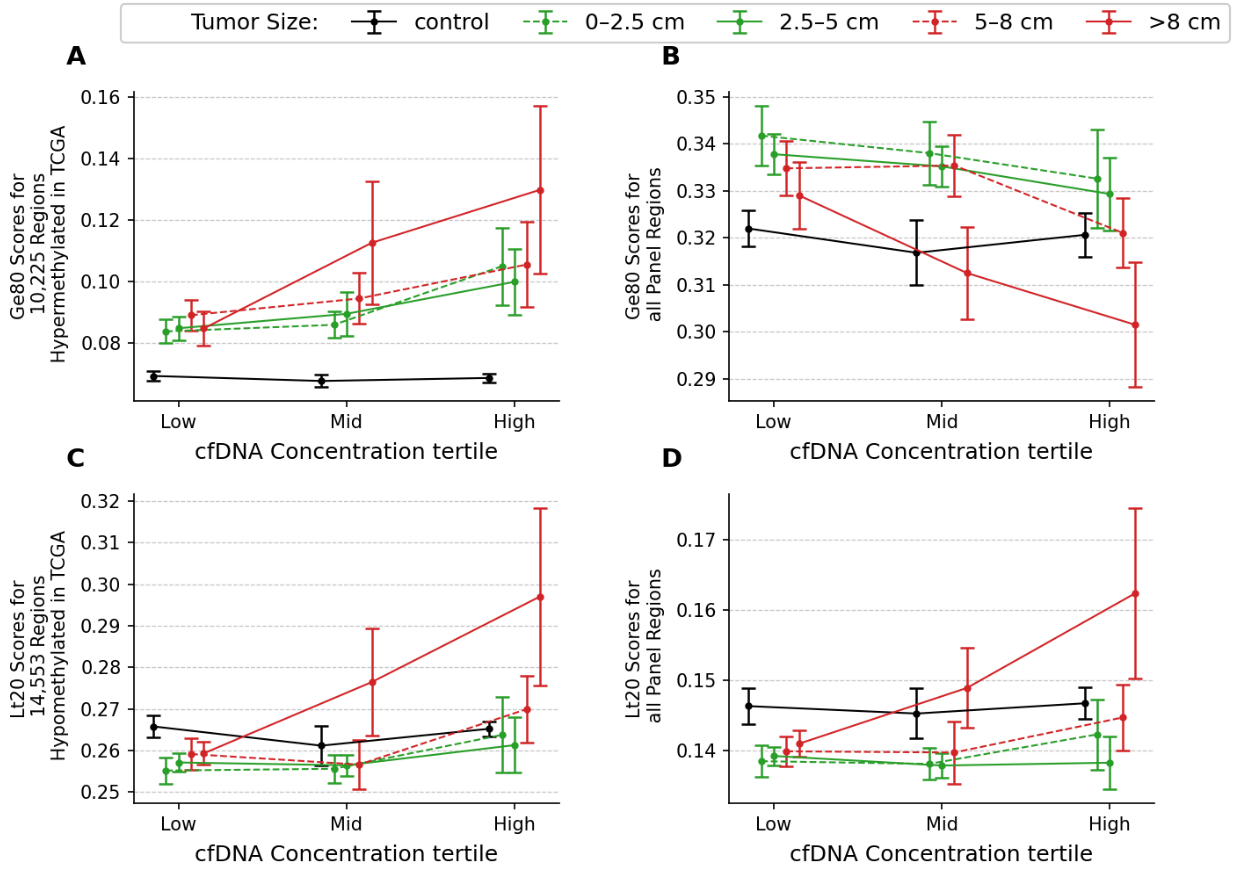


**Supplementary Figure 4. Variation of hypermethylation and hypomethylation scores with cfDNA concentration in our main cohort, stratified by tumor size and further by cfDNA concentration tertiles within each size group.** Each plot shows mean methylation scores for a chosen metric with 95% confidence intervals (Wald). Tumor size groups were defined as 0–2.5 cm, 2.5–5 cm, 5-8 cm and >8 cm; controls are shown separately. (A) Mean cfDNA Ge80 hypermethylation scores for 10,225 regions identified as hypermethylated in TCGA tumor tissue. (B) Mean cfDNA Ge80 hypermethylation scores across all 551,803 panel regions. (C) Mean cfDNA Lt20 hypomethylation scores for 14,553 regions identified as hypomethylated in TCGA tumor tissue. (D) Mean cfDNA Lt20 hypomethylation scores across all 551,803 panel regions.

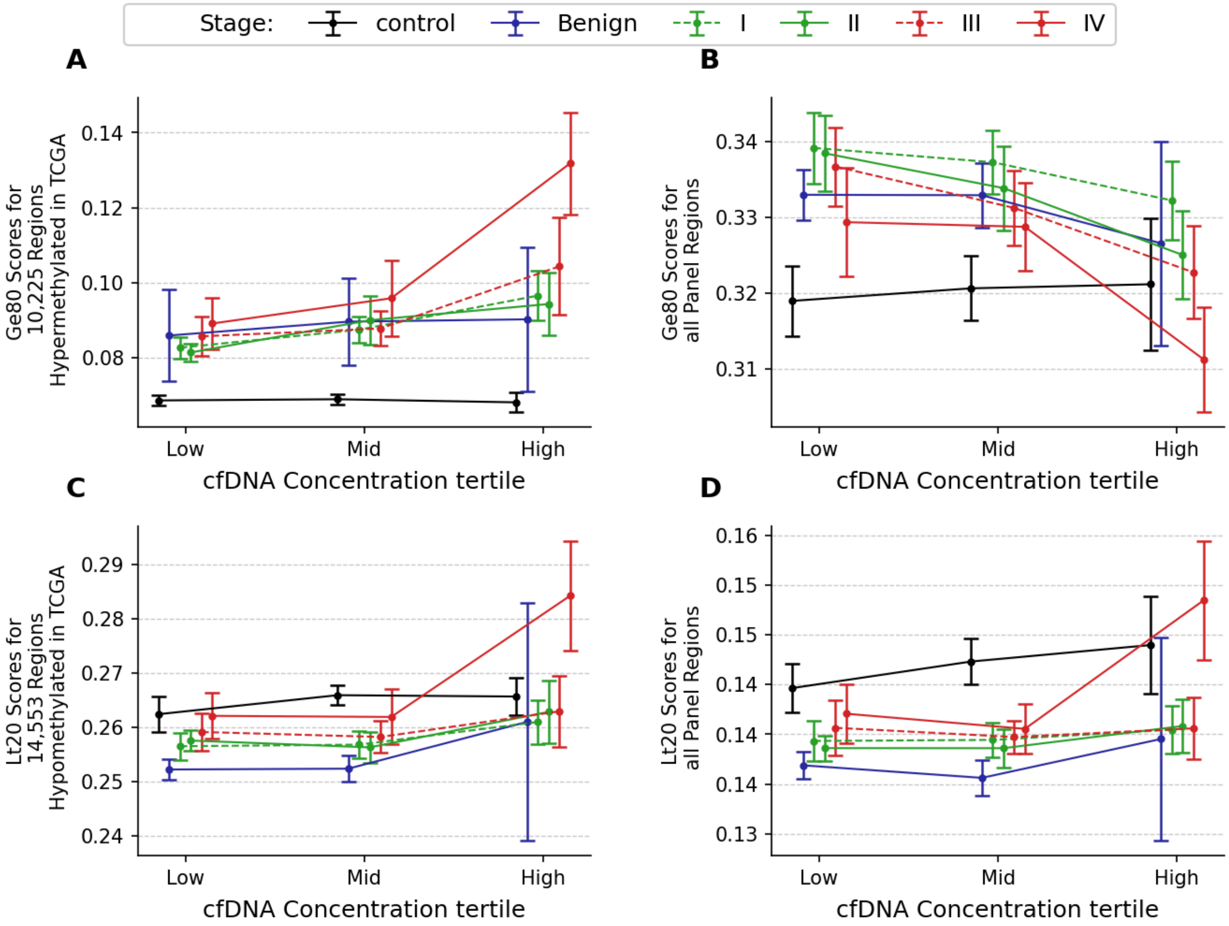


**Supplementary Figure 5. Variation of hypermethylation and hypomethylation scores with cfDNA concentration in the main cohort, stratified by stage and by cfDNA concentration tertiles defined globally.** Each plot shows the mean and 95% CI (Wald) for a chosen metric, stratified by stage-concentration tertile combinations. Concentration tertiles were defined globally across stages. The metrics in the depiction are: **(A)** Mean cfDNA Ge80 hypermethylation scores for 10,225 hypermethylated regions identified from TCGA analysis. **(B)** Mean cfDNA Ge80 hypermethylation scores for all 550K panel regions. **(C)** Mean cfDNA Lt20 hypomethylation scores for 14,553 hypomethylated regions identified from TCGA analysis. **(D)** Mean cfDNA Lt20 hypomethylation scores for all 550K panel regions.

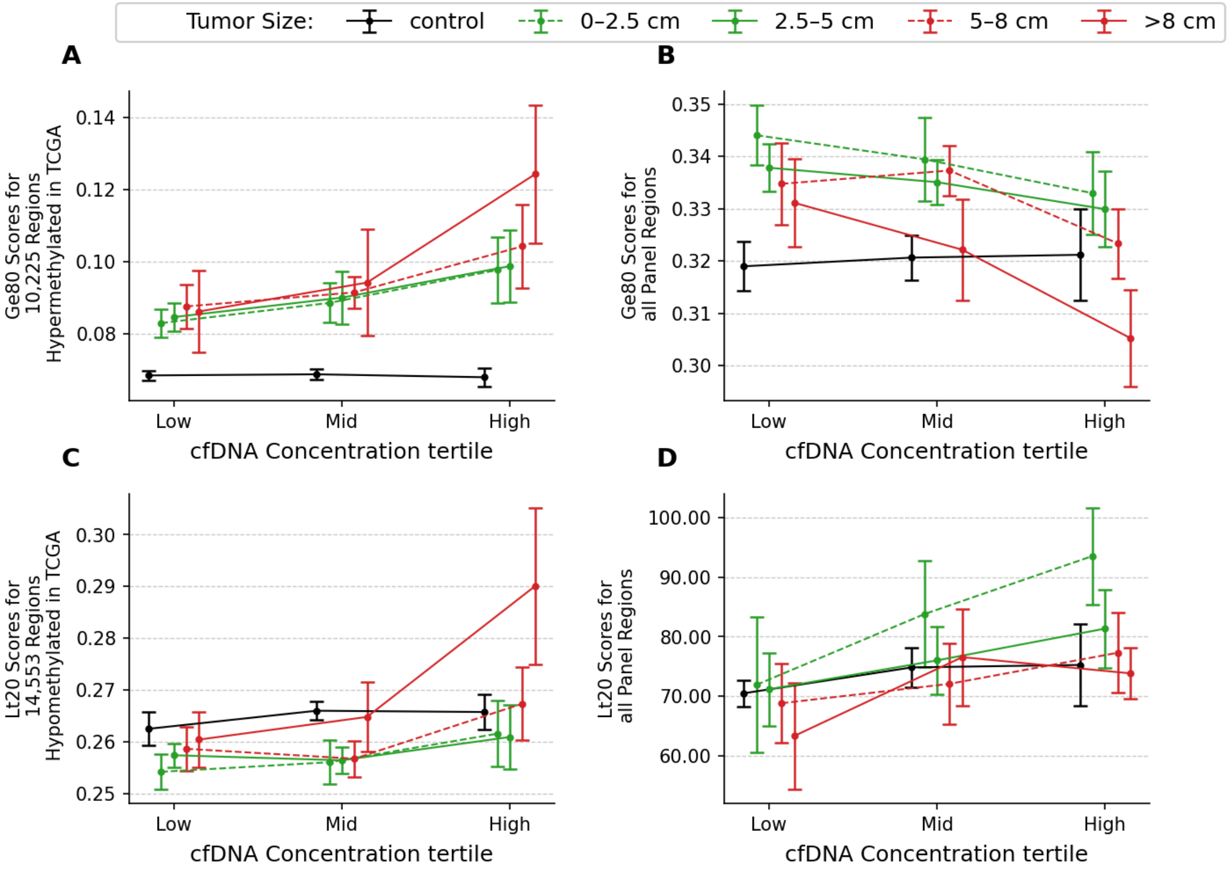


**Supplementary Figure 6. Variation of hypermethylation and hypomethylation scores with cfDNA concentration in our main cohort, stratified by tumor size and by cfDNA concentration tertiles defined across size-groups.** Each plot shows mean methylation scores with 95% confidence intervals (Wald). Tumor size groups were defined as 0–2.5 cm, 2.5–5 cm, 5-8 cm and >8 cm; controls are shown separately. (A) Mean cfDNA Ge80 hypermethylation scores for 10,225 regions identified as hypermethylated in TCGA tumor tissue. (B) Mean cfDNA Ge80 hypermethylation scores across all 551,803 panel regions. (C) Mean cfDNA Lt20 hypomethylation scores for 14,553 regions identified as hypomethylated in TCGA tumor tissue. (D) Mean cfDNA Lt20 hypomethylation scores across all 551,803 panel regions.

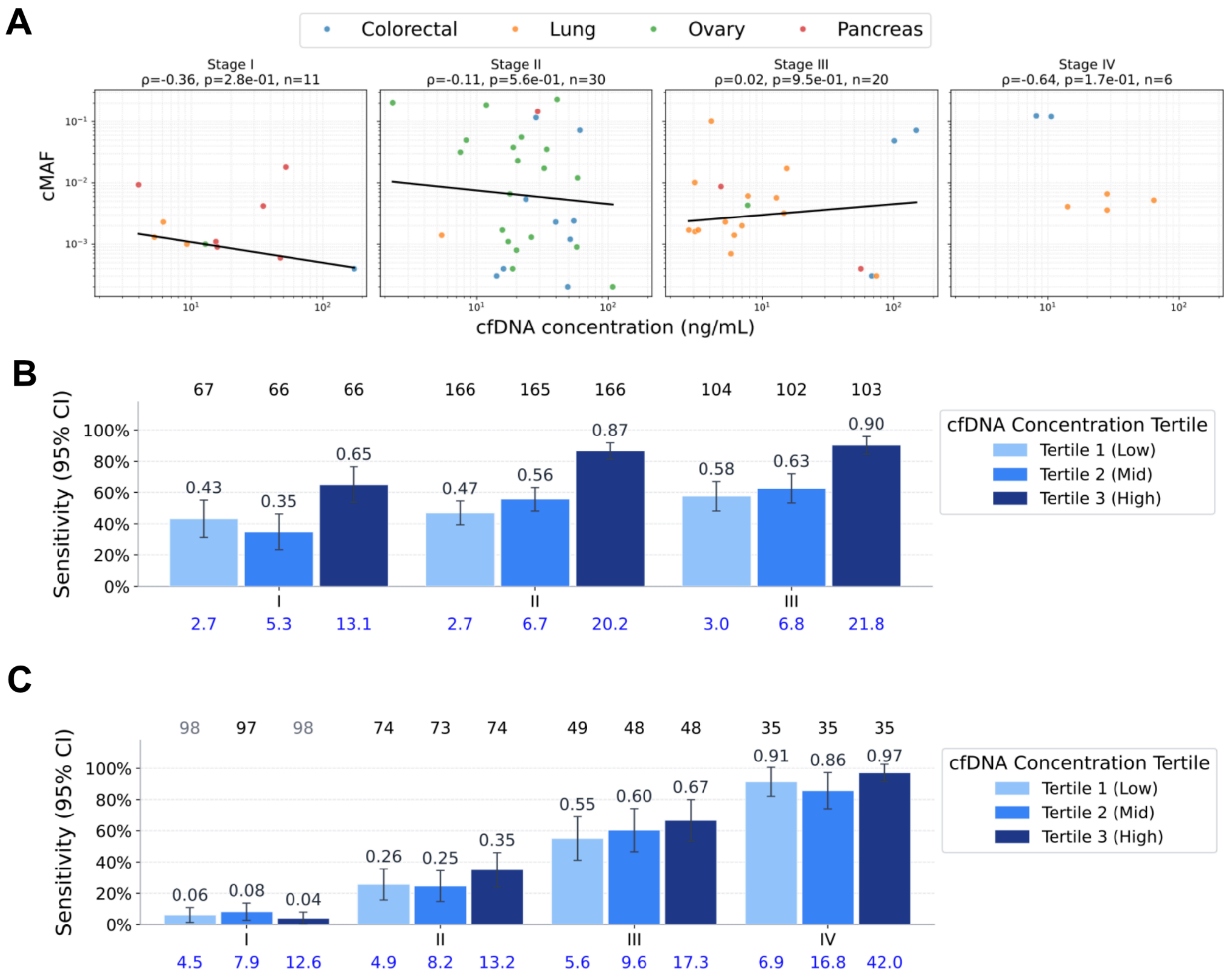


**Supplementary Figure 7. The association between cfDNA concentration, circulating Mutation Allele Frequency (cMAF), and detection sensitivity in published datasets. (A)** Circulating mutation allele frequency for confirmed somatic variants vs cfDNA concentration from Mattox et al. Cancer Discov. 2023**,** stratified by stage. Plots show the Theil-Sen regressor line along with the Spearman coefficient, p value and number of samples. Cancer types in each plot are color coded. **(B)** Classifier sensitivity with 95% CI (Wald) for concentration tertiles within each stage, shown as bar plots, for the Cohen et al. Science. 2018 dataset. The number of samples in each tertile within each stage is indicated above the plot. The median concentration value for each tertile-stage combination is indicated in blue below the bar. **(C)** Same as (B) but for the Bredno et al. PLoS One. 2021 dataset.

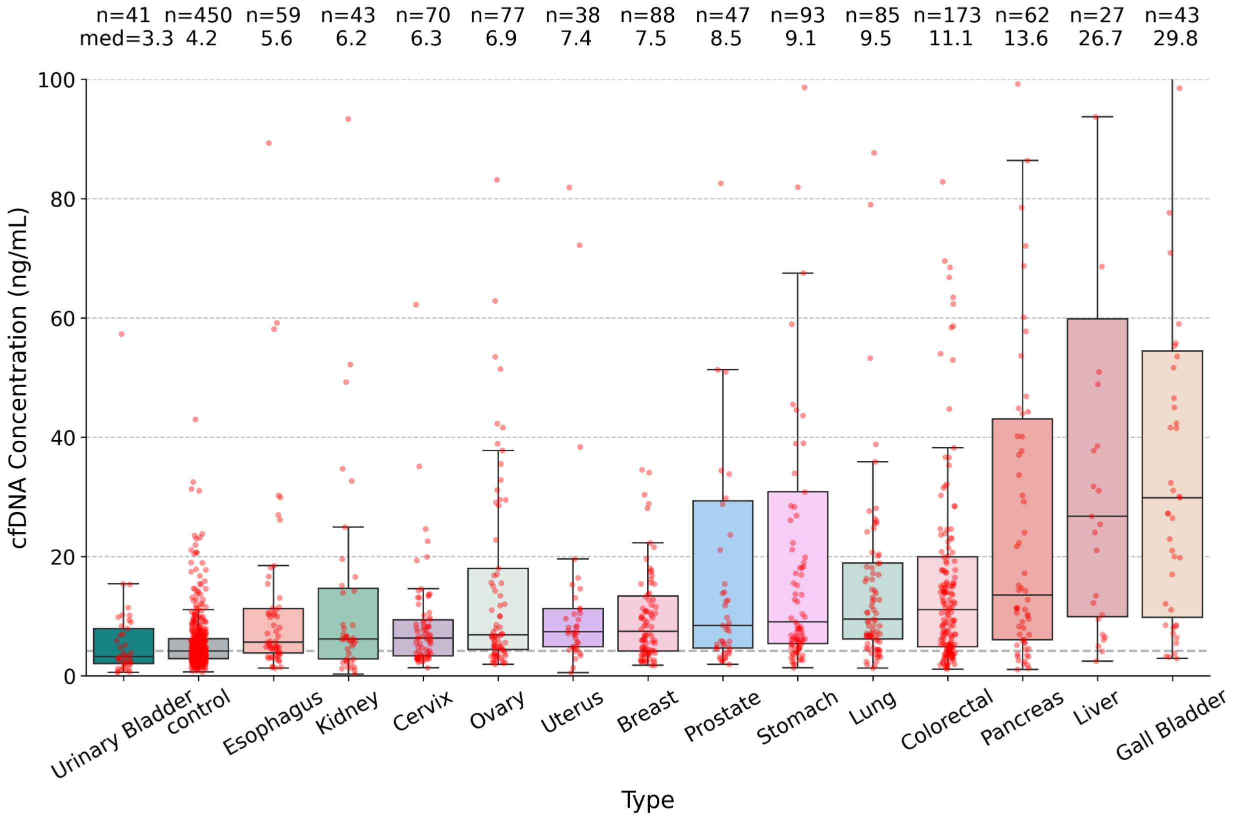


**Supplementary Figure 8. Variation of cfDNA concentration with cancer type in the main cohort.** Violin plots show the distribution of cfDNA concentration for controls and for each cancer type. For each cancer type, median concentration values and sample size are indicated at the top.

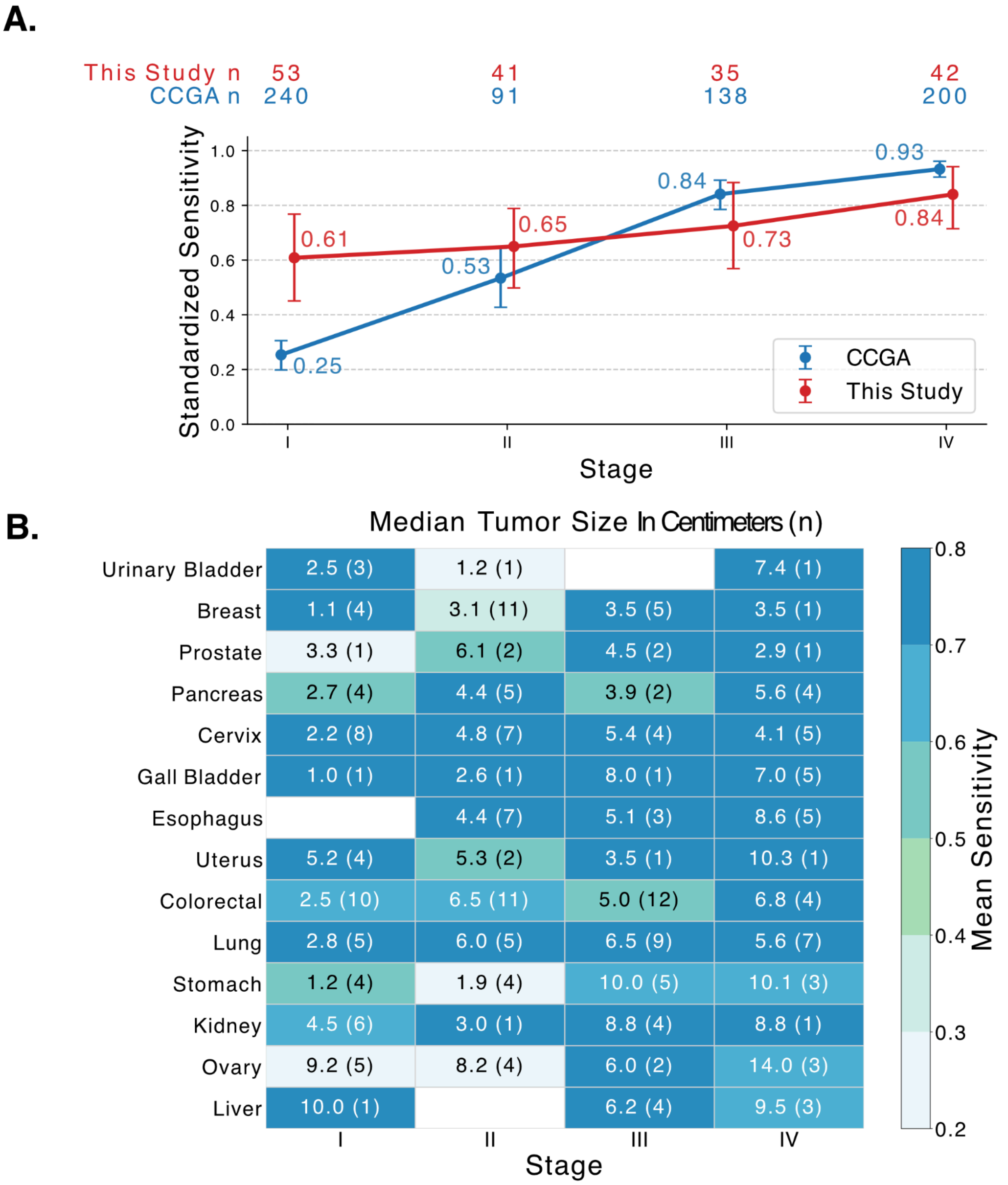


**Supplementary Figure 9. Comparison with CCGA. (A)** Independent validation set sensitivities of this study and the CCGA study, after standardizing cancer type and stage distributions, and restricting analysis to 9 non-screenable cancers that were diagnosed upon clinical presentation in both cohorts (esophagus, gall bladder, liver, stomach, ovary, uterus, urinary bladder, kidney, pancreas). Sample counts from each study are indicated at the top. **(B)** Median tumor sizes in cm by cancer type and stage for the independent validation set. Cells are color coded by detection sensitivity. Numbers in round brackets indicate sample counts. Tumor sizes were not available for 122/337 cases.

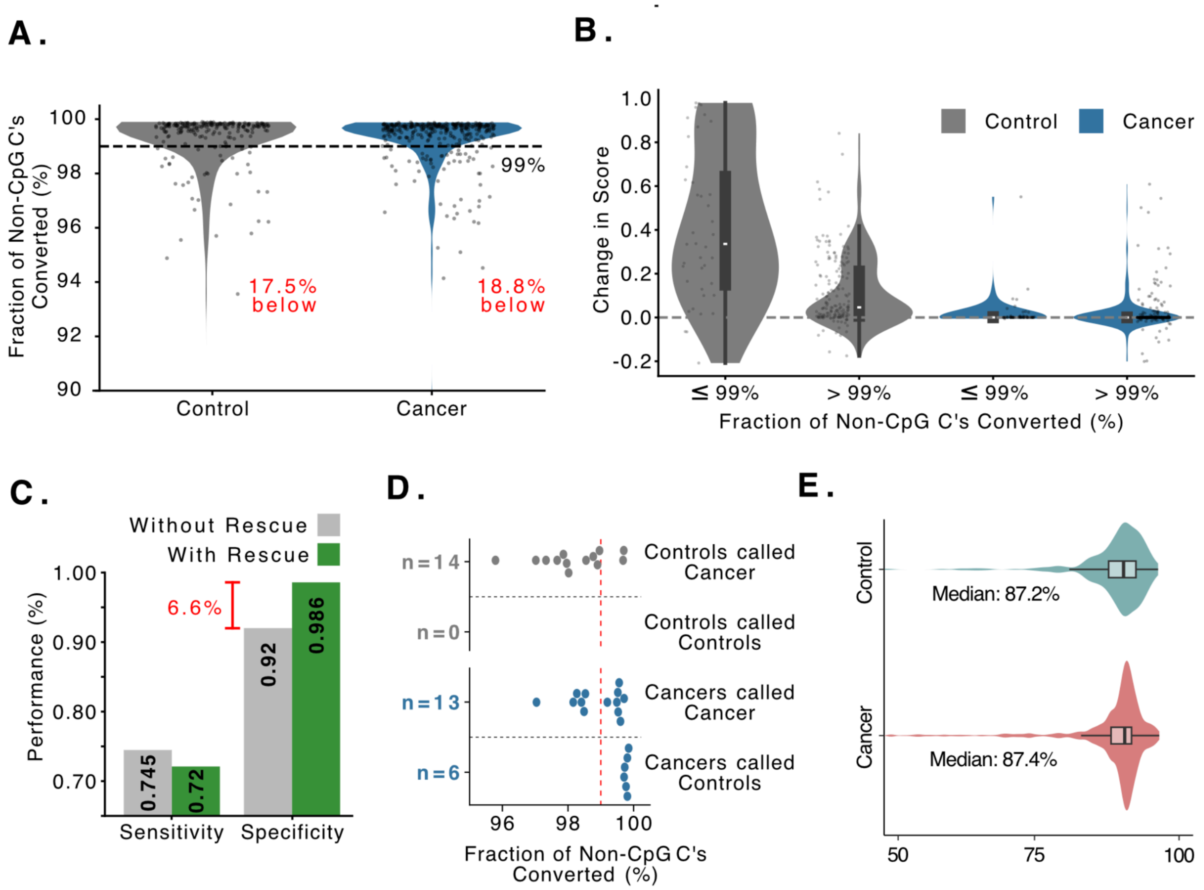


**Supplementary Figure 10. Effects of rescue step on the independent validation cohort.** (**A)** Violin plots show distributions of the fraction of non-CpG cytosines converted, stratified by control and cancer status. Each dot represents a sample. The dashed line marks the 99% conversion efficiency cutoff; the proportion of samples below this threshold is indicated for each group. (**B)** Violin plots show increase in classifier score when the rescue step is omitted, stratified by cancer and control status, and further by whether the fraction of non-CpG cytosines converted is <99% or not. Each dot represents a sample. (**C)** Bar plot shows sensitivity and specificity with and without rescue. The drop in specificity upon skipping rescue of ~6% is highlighted, even at a slight gain in sensitivity. **(D)** Dot plots show samples whose prediction class changes when rescue is omitted, stratified by control and cancer status, and further by the new predicted class. The red vertical line represents the 99% cutoff for the fraction of non-CpG cytosines converted. The drop in specificity and slight rise in sensitivity when rescue is omitted is largely due to samples below this cutoff. (**E)** Violin plots for cases and controls separately show that there is no bias in the number of reads filtered by this step between cases and controls and that a median of ~87% of reads are retained, although a very small fraction of samples lose ~50% of reads.

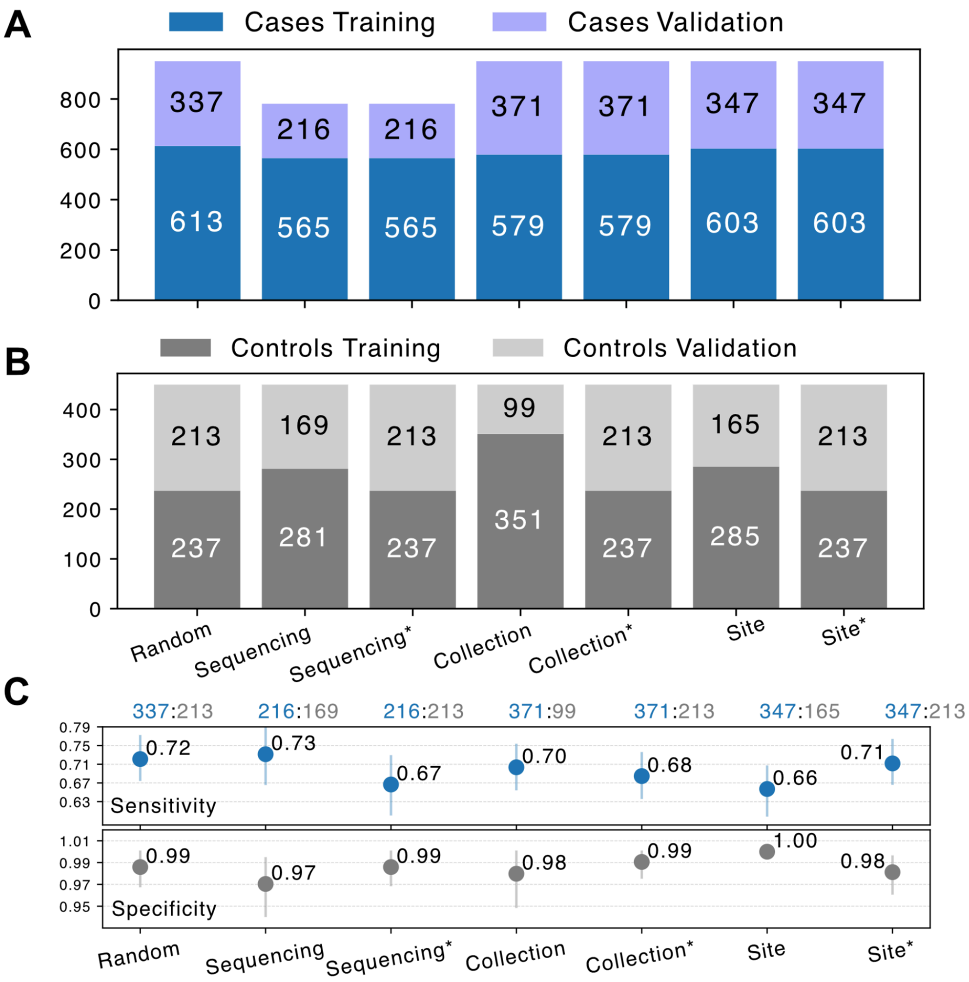


**Supplementary Figure 11: Robustness against multiple training-validation splits. (A, B)** Bar plots show the number of cases and controls, respectively, in each of the splits into training and independent validation; non-random splits are defined by a chronological cutoff for collection time and for sequencing batch splits, and by site number order for site-wise splits (with one exception: controls were recruited at 3 sites 26, 27 and 35; site 27 was considered in the training set since it contributed ~58% of all controls, and sites 26 and 35 in the validation set). Site numbers were assigned arbitrarily at the project outset. *Splits marked by * have controls randomized across the split.* Sequencing splits have fewer samples in validation because only 10/14 cancer types are represented adequately in training**. (C)** Dot plots show independent validation set sensitivity and specificity for classifiers derived from the various adversarial splits. Error bars show 95% CIs (bootstrap n=1000). Numbers at the top are counts of cases and controls in the independent validation set.

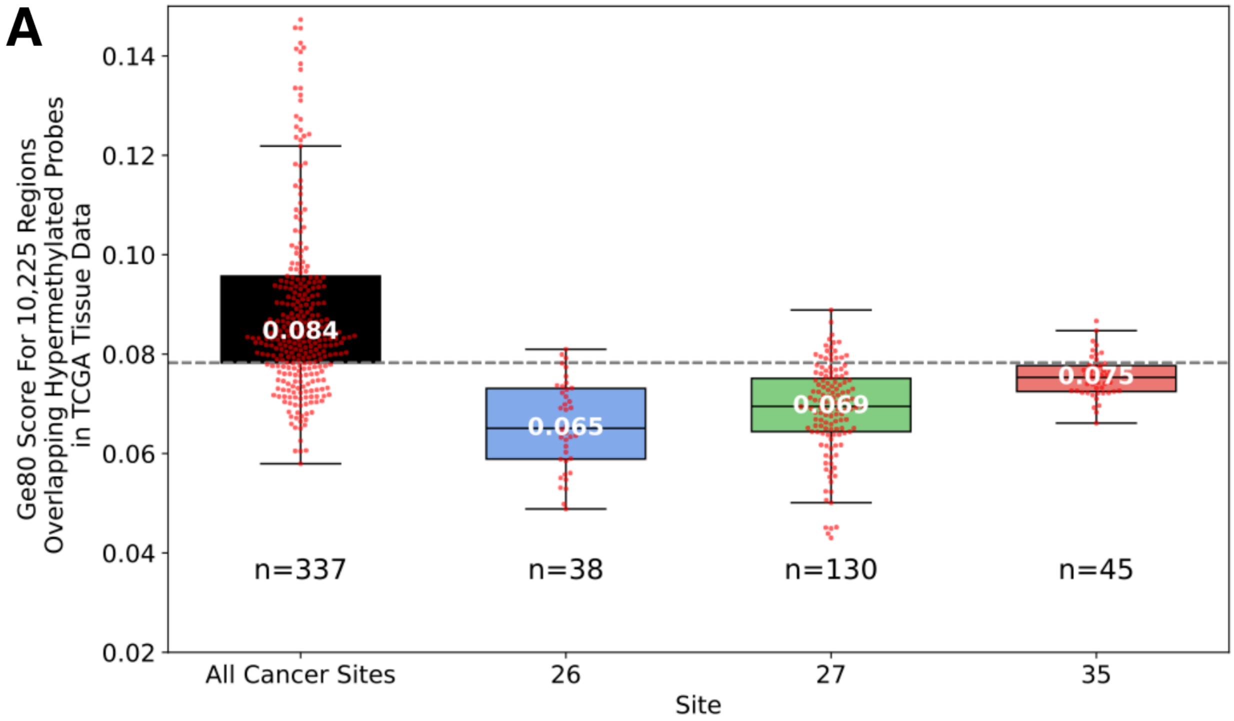


**Supplementary Figure 12. Ge80 methylation scores in TCGA-defined cancer-associated regions in cases and in controls stratified by recruitment site**. Box plots show mean Ge80 hypermethylation scores across 10,225 TCGA hypermethylated regions for controls at sites 26 (n=38), 27 (n=130), and 35 (n=45), compared against all cancer cases pooled across 40 sites (n=337). The dashed line indicates the first quartile score across all cancer cases.

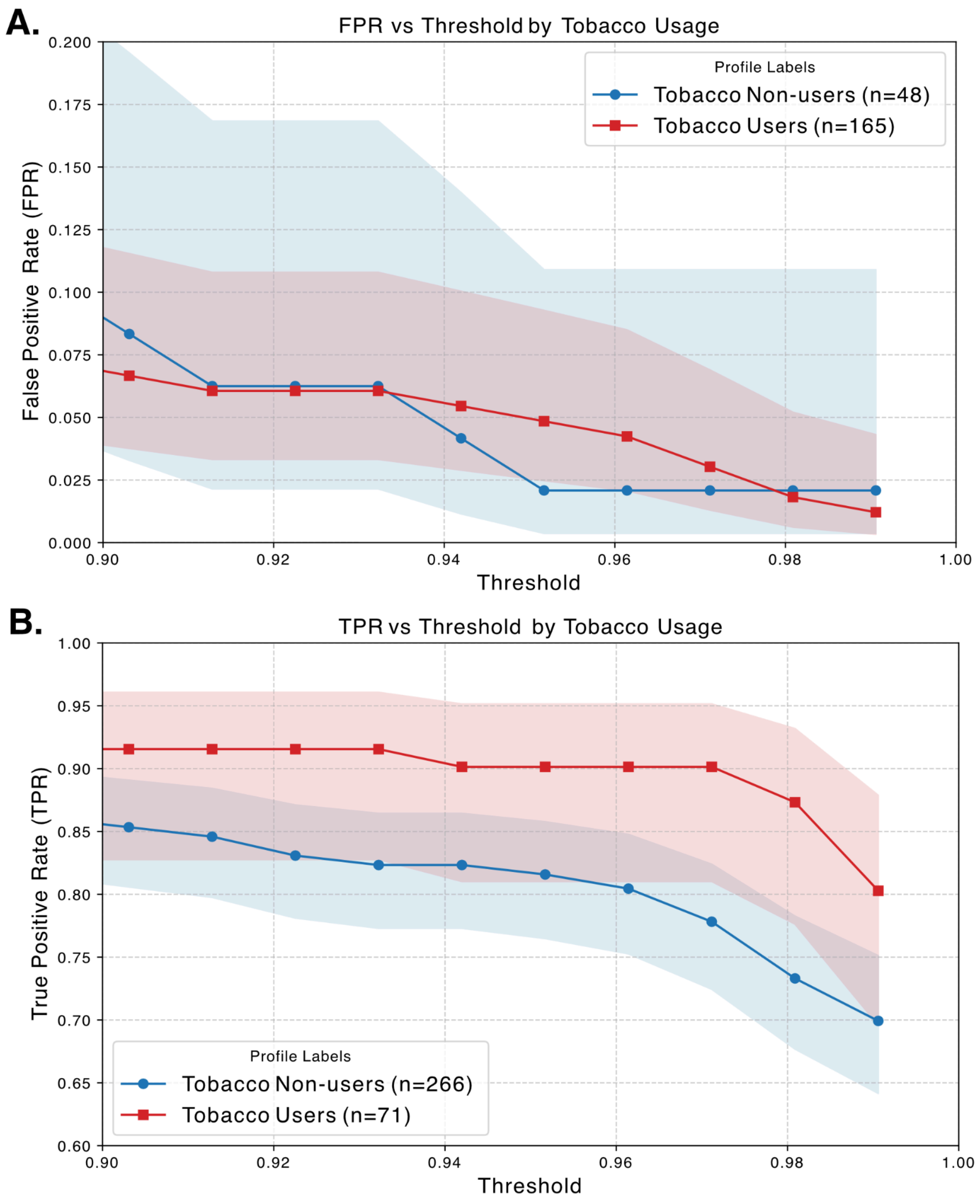


**Supplementary Figure 13. Classifier performance in independent validation stratified by tobacco use status. (A)** False positive rate (1-specificity), and **(B)** True positive rate (sensitivity), evaluated across classification thresholds from 0.90 to 0.9906 for tobacco users and non-users separately, with 95% confidence intervals (Wilson) shown as shaded bands. 0.9906 is the cutoff for positive calls.

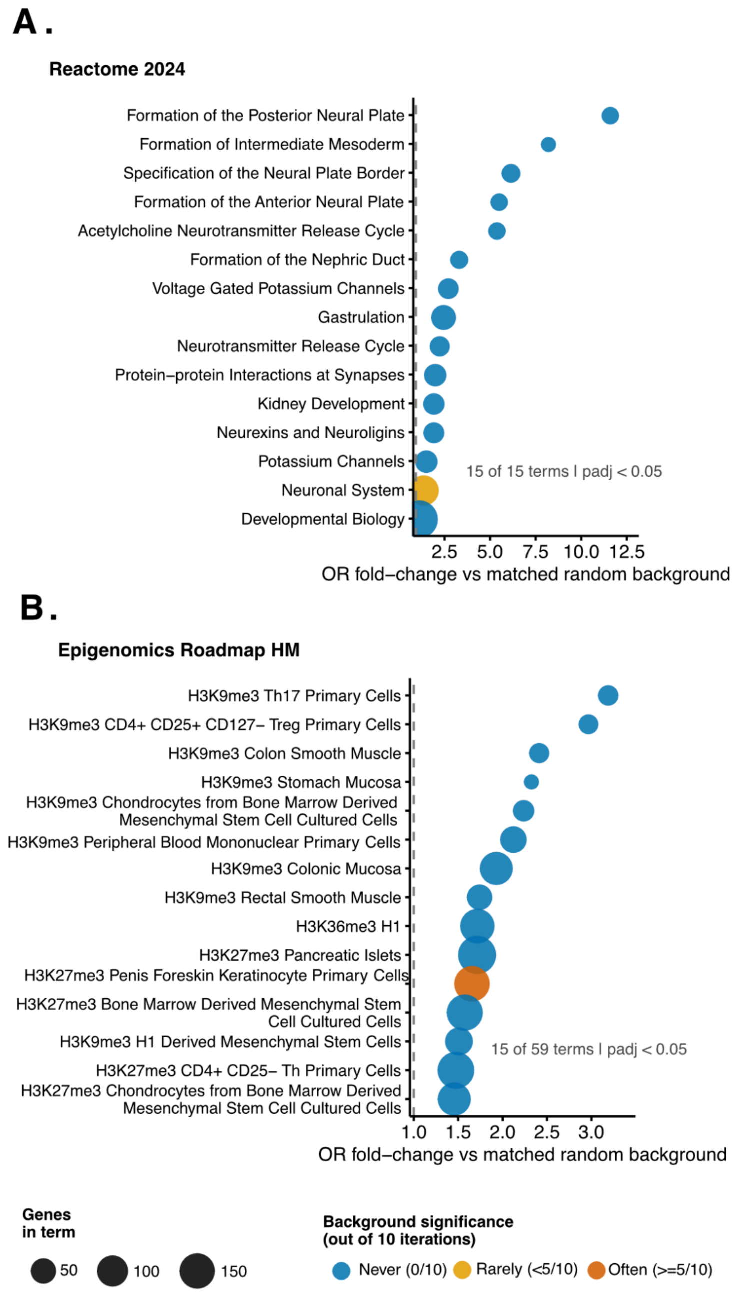


**Supplementary Figure 14. Over-representation analysis (ORA) for promoter genes associated with classifier regions against 10 matched random background gene sets.** Results are shown for 2 databases with at least one significant term (adjusted p < 0.05): **(A)** Reactome 2024: All 15 significant terms shown. **(B)** Epigenomics Roadmap HM ChIP-seq: 15 of 59 significant terms shown. The x-axis shows the odds ratio fold-change for classifier regions against ten matched background iterations. Dot size represents the number of promoter genes in the term. Color denotes background specificity.

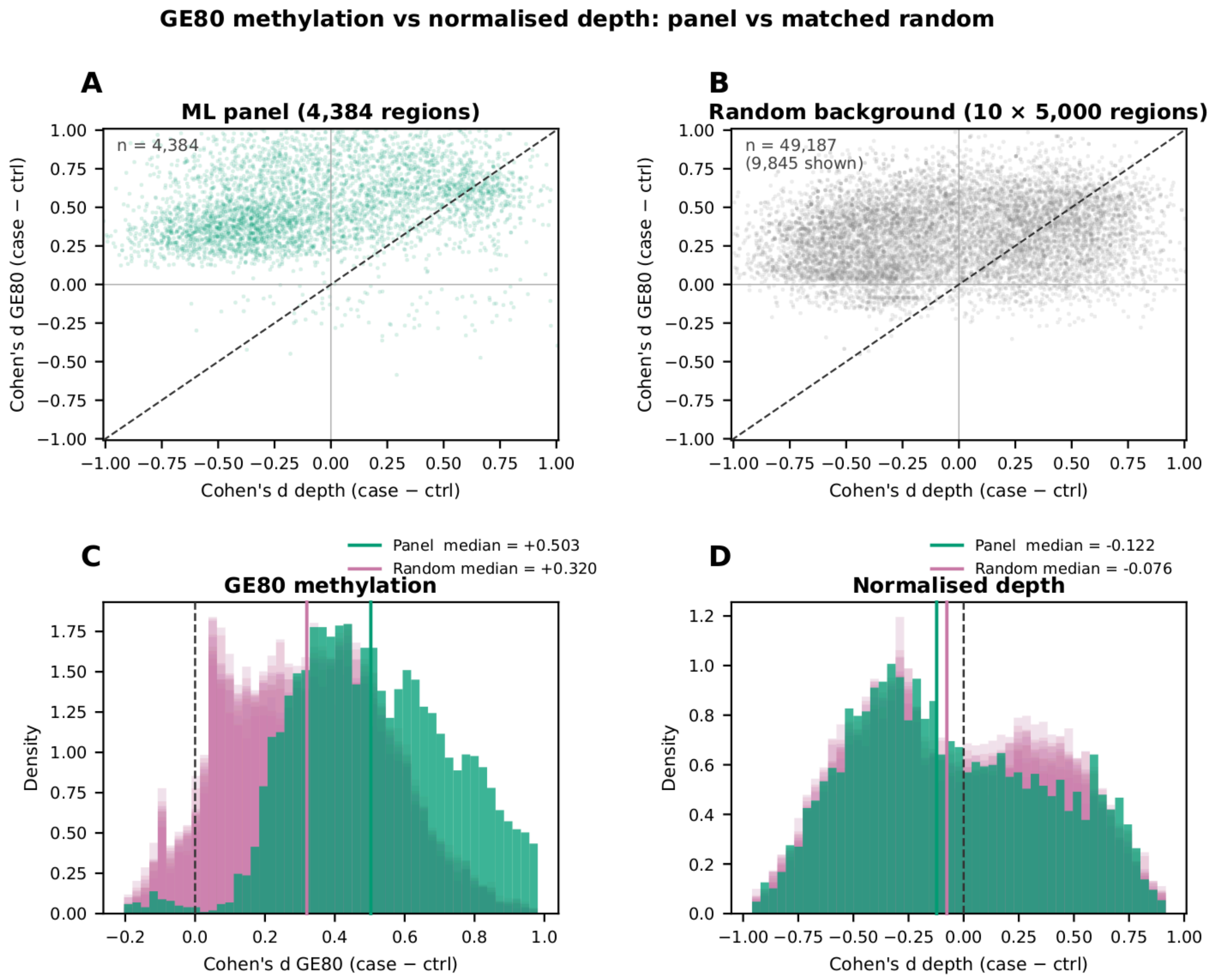


**Supplementary Figure 15. Discriminative capability of methylation vs fragment depth. (A)** Cohen's d (case − control) for Ge80 hypermethylation scores (y-axis) versus normalised fragment depth (x-axis) for classifier regions. **(B)** Cohen's d (case − control) for Ge80 hypermethylation scores (y-axis) versus normalised fragment depth (x-axis) for background regions (n = 49,187 across 10 iterations). **(C)** Distribution of Cohen's d for Ge80 hypermethylation scores. **(D)** Distribution of Cohen's d for normalised fragment depth.

### SUPPLEMENTARY METHODS

#### 1. Ethics Approval & Consent to Participate

In the EMERGE study (Epigenetic Markers for Early and Robust Cancer Detection in a General Population), the names of the Ethics Committees that approved the study and their respective registration numbers appear below. In total, 22 IEC approvals were obtained covering 43 recruitment sites. Informed consent was obtained from all individual participants included in the study.

1. Ethiclin Pvt Ltd IEC: ECR/350/Indt/KA/2021
2. HCG-BBHIO Ethics Committee: ECR/994/INST/KA/2017/RR-21
3. BGS Global Hospitals IEC: ECR/128/Inst/KA/2013/RR-19
4. Prakriya Hospitals IEC: ECR/1412/Inst/KA/2020
5. Shreyas Hospital IEC: ECR/962/Inst/MH/2017/RR-20
6. Shanmuga Medical Research Foundation Trust IEC: ECR/1046/Inst/TN/2018/RR-21
7. Erode Cancer Centre IEC: ECR/319/INST/TN/2013/RR-19
8. CARE Multispeciality Hospitals IEC: ECR/94/Inst/AP/2013/RR-21
9. Shrey Hospital IEC: ECR/1302/Inst/GJ/2019
10. HP Poddar Memorial Clinic and Nursing Home IEC: ECR/1555/Inst/WB/2021
11. Manavata Research Center IEC: ECR/500/Inst/MH/2013/RR-20
12. Amravati Ethics Committee, SSCHACF: ECR/432/Inst/MH/2013/RR-19
13. Galaxy Care Multispeciality Hospital IEC: ECR/1379/Inst/MH/2020
14. Aurangabad Health Care and Research LLP IEC: ECR/325/Indt/MH/2020
15. Telerad RxDx Healthcare Pvt Ltd IEC: ECR/1494/Inst/KA/2021
16. HCG-Central Ethics Committee: ECR/386/Inst/KA/2013/RR-19
17. Sangini Hospital IEC: ECR/147/Inst/GJ/2013/RR-19
18. Krishna Institute of Medical Sciences "Deemed to be University" IEC: ECR/307/Inst/MH/2013/RR
19. Shri Siddhivinayak Ganapati Cancer Hospital IEC: ECR/588/Inst/MH/2014/RR-21
20. Chittaranjan National Cancer Institute IEC: EC/NEW/INST/2020/946
21. Saveetha Medical College and Hospital IEC: ECR/724/Inst/TN/2015/RR-24
22. Central IEC: ECR/390/Indt/MH/2024

#### 2. Collection

Whole blood was collected from eligible subjects after informed consent was obtained. For subjects with benign, precancerous or malignant lesions, blood was collected (2 × 10 mL) at the time of diagnosis before initiation of any cancer-directed therapy. For healthy controls, blood was collected similarly but at the time of recruitment. All blood samples were collected in Streck tubes (Streck, 218962), transported to the central laboratory of Strand Life Sciences at Bengaluru, India within 72 hours and immediately fractionated to isolate plasma and buffy coat, which were stored at –80°C until further analysis.

#### 3. Sample Processing

Peripheral whole blood collected in Streck tubes was processed according to a standardized two-step centrifugation protocol to isolate plasma. Briefly, blood was centrifuged at 1900 × g for 22 minutes at 4°C, and the resulting supernatant was transferred to 15 mL tubes and subjected to a second centrifugation at 3200 × g for 20 minutes at 4°C. The resulting cell-free plasma was extracted and aliquoted into 4 mL portions, which were stored at −80 °C until further processing. The buffy coat was separated, preserved in RNAlater (Thermo Fisher Scientific, AM7021), and stored at −80°C.

Cell-free DNA (cfDNA) was extracted from 4 mL of plasma using the Apostle MiniMax High Efficiency cfDNA Isolation Kit (Apostle Bio, A17622-384) following the manufacturer's protocol. cfDNA concentration was quantified on a Qubit 4.0 fluorometer, using either Qubit High Sensitivity dsDNA Assay (Invitrogen, Q32854), or LeoNext 1x dsDNA HS Assay Kit (Genes2Me, NGS3203-02) and fragment size distribution was assessed on the Agilent 4200 TapeStation (RRID:SCR_018435) with High Sensitivity D1000 ScreenTapes (Agilent, 5067-5584). Up to 20 ng of total cfDNA were taken for library preparation.

Libraries for methylation sequencing were generated using the NEBNext Enzymatic Methyl-Seq Kit (Twist Biosciences, 101977), according to the manufacturer's instructions. In brief, up to 20 ng of cfDNA was spiked with 0.02 ng unmethylated lambda DNA and 0.001 ng methylated pUC19 controls, and subjected to end-repair, A-tailing, and ligation of indexed adapters. Protection of 5-methylcytosine (5mC) and 5-hydroxymethylcytosine (5hmC) was achieved with TET2 and T4-BGT enzymes prior to APOBEC3A-mediated deamination of unprotected cytosines. Following a bead-based cleanup and a safe-stop incubation at −20°C for ≥48 hours, libraries were PCR amplified with cycle number adjusted to input amount (e.g., 11 cycles for 10–14 ng, 10 cycles for 15–20 ng). Unique dual indices were incorporated during amplification. Library yield was quantified by Qubit (Invitrogen, Q32854, RRID:SCR_026883) and size distribution assessed on the TapeStation 4200 (Agilent G2991BA, RRID:SCR_018435); libraries exhibiting ≥400 ng total yield and an expected size profile (primary peak ~350 bp, secondary peak ~500 bp) were carried forward.

#### 4. Capture Panel & Sequencing

The 123 Mb Twist Human Methylome Panel (v2022-06, ~551,803 regions, ~123 Mb, ~3.98 million CpGs) served as our primary discovery tool. It targets approximately 84% of the CpG islands in the Human Genome along with a selection of the surrounding shores, shelves, and open sea regions. These CpG island regions are biologically significant in oncology as they typically remain unmethylated to keep housekeeping genes active, but frequently undergo widespread epigenetic remodeling and become aberrantly hypermethylated during tumorigenesis.

Targeted hybridization capture was performed overnight using the above panel. Libraries were pooled in sets of eight (187.5 ng per library) or twenty-four (166.7 ng per library) prior to capture. Post-capture enrichment was performed by PCR (7 cycles), and final libraries were quantified by

Qubit (Invitrogen, Q32854) and assessed on the TapeStation 4200 (Agilent G2991BA). Typical post-capture libraries had concentrations of ~25–35 ng/µL and average fragment sizes of ~350 bp.

Sequencing was performed with paired-end 2×150 bp reads on Illumina platforms (NovaSeq 6000, RRID:SCR_016387 S4 Reagent Kit v1.5 (300 cycles) (Illumina, 20028312) or NovaSeq™ X Series (RRID:SCR_024568) 25B Reagent Kit (300 Cyc) (Illumina, 20104706) or NovaSeq X Series 10B Reagent Kit (300 Cycle) (Illumina, 20085594), targeting 62.5 Gb of raw data per sample.

##### 5. Bioinformatics Processing

Raw sequencing data were demultiplexed using bcl-convert v1.0.0 (Illumina) to generate per-sample FASTQ files. Reads were processed through our in-house pipeline FrAnaTk (Fragment Analysis Toolkit), orchestrated with NextFlow to ensure reproducibility and modular execution of the steps below.

Adapter sequences and low-quality bases were removed using BBduk v38.96 (SourceForge) with the following parameters: ktrim=r, k=19, mink=5, hdist=1, hdist2=0, qtrim=r, trimq=14, and minlength=36. Pre- and post-trimming quality was assessed using FastQC v0.12.1 (Babraham Bioinformatics).

Trimmed reads were aligned with BWAMeth v0.2.5, which accommodates APOBEC3A-induced deamination of unprotected cytosines by aligning to two reference sets of the human genome (hg38, UCSC): one is the original and the second is pre-converted. Additionally, a panel of control genomes including Lambda (J02459.1) and pUC19 (L09137.2) was used as controls to measure deamination and protection efficiency. Further HPV16 (NC_001526.4), and HPV18 (NC_001357.1) were used to identify reads sourced from the HPV genomes, which may be indicative of cervical carcinoma. Duplicate reads were marked using Samblaster v0.1.26, and the resulting BAM file was sorted and indexed with Samtools v1.14. This sorted, indexed BAM file was used as the final alignment output for all downstream analyses.

Methylation calling: To determine conversion efficiency, MethylDackel v0.6.1 (extract and merge context modules) was run on the sorted, indexed BAM file, generating bedGraph files that record counts of cytosines and thymines at each CpG, CHG, and CHH context genome-wide, from which pConvNonCpG and pConvLambda, indications of conversion efficiency, were calculated.

Quality control metrics: Quality control (QC) metrics were generated from the sorted, indexed BAM file using multiple software tools and custom scripts to characterize library and sequencing performance (Samtools -f 64, -f 128 and -F 4 for on target alignment, Picard v2.18.29, and custom code implemented in R to compute coverage from Picard output and methylation metrics like pMeth and pConvNonCpG from MethylDackel outputs). The programming languages used were R (v4.4.3) and Python (v3.12.2).

Fragment-level methylation profiling: A customized version of wgbs-tools was employed to generate a PATR file from the above sorted indexed BAM file. Again, Samtools filtering was applied with flags -f 3 -F 3852 to retain only properly paired, high-quality reads and to exclude unaligned, duplicate, supplementary, or otherwise flagged alignments. In this process, both mates of each read pair were merged into a single fragment, fragments that did not overlap any CpG sites were excluded, and the remaining fragments were represented one per line with a “C” or “T” for each overlapping CpG. Unlike a PAT file, this PATR file is unclustered, i.e., fragments with the same methylation pattern are retained individually and not clustered together into a single pattern.

HPV Score: The fraction of aligned reads mapped to the HPV16 (NC_001526.4) and HPV18 (NC_001357.1) genomes multiplied by 1 million was reported as the HPV score.

##### 6. Conversion Assessment & Fragment-Level Rescue

To ensure accurate methylation calling and minimize biases arising from incomplete cytosine deamination, we implemented a custom fragment conversion quality assessment procedure tailored to our APOBEC3A-based conversion workflow. For each fragment represented in the PATR files, we quantified the extent of APOBEC-mediated C→T conversion by examining bases aligned to non-CpG cytosines (C) and the complementary guanines (G) in the reference genome, positions where near-complete conversion is expected for unmodified cytosines following APOBEC-induced deamination.

For downstream analyses, only fragments with a conversion fraction ≥ 95% (i.e., ≥ 95% of non-CpG cytosines converted to thymine) were retained, and fragments failing to meet this threshold were excluded in the rescue step. This custom filtering step was used to prevent artifacts from incomplete conversion from biasing both methylation and fragmentomic metrics.

We calculated pConvNonCpG, the sample-level non-CpG conversion fraction, as the total number of reads with thymine substitutions across all CHG and CHH reference sites divided by the total number of reads at those sites. pConvCpGLambda, the conversion fraction observed in the lambda control sequences, was also assessed as an alternative measure of conversion efficiency.

##### 7. Region-Level Methylation Feature Derivation

Fragment bed and PATR files generated from the bioinformatics pipeline were further processed to compute region-level methylation features for each of the 551,803 targeted regions in the Twist Human Methylome panel. Only fragments that contained at least three aligned reference CpGs and had at least one CpG overlapping a target region were included in feature calculations to ensure reliable locus-specific signals.

For each target region in each sample, we computed the following metrics: 1) Ge80: the fraction of fragments mapping to the region that exhibited ≥80% of CpG sites methylated, representing regions with high methylation enrichment; 2) Lt20: the fraction of fragments mapping to the region that exhibited <20% of CpG sites methylated, representing regions with low methylation levels.

These region-wise, sample-wise scores were assembled into a feature matrix and used as input for subsequent classifier training and evaluation.

#### 8. Fragmentomics Feature Derivation

Arm-Level Fragment Size Distribution (Bao et al. *Nature Med.* 2025): The genome was divided into 39 autosomal chromosomal arms. For each chromosomal arm, the proportion of fragments in each of the size bins 100-104 bp, 105-109 bp, ... , 215-219 bp was computed. This resulted in 39 x 24 = 936 features for each sample .

Region-Level Short and Long Fragment Coverage (Bao et al. *Nature Med.* 2025): For each region in the capture panel, the coverages of short (65-150 bp) fragments and long (151-220 bp) fragments were computed and corrected to the GC content of the region. The autosomal genome was divided into non-overlapping 5Mb bins, resulting in 495 bins. In each bin, the coverages of any overlapping panel regions were summed up, resulting in 495 short fragment coverages and 495 long fragment coverages for each sample.

The DELFI Score (Cristiano et al. *Nature* 2019): The ratio of short (65–150 bp) to long (151–220 bp) fragments in each of the 495 5 Mb bins were computed.

Fragment End Motif Fraction (Jian et al. *Cancer Disc.*, 2020): For each sample, the proportion of 4-mer end motifs at the 5' end were computed. This resulted in 256 end motif frequencies for each sample.

Nucleosome Protection Score (Erger at al. *Genome Med*. 2020): Nucleosome positioning was assessed across a targeted subset of approximately 20,000 transcription start sites (TSS) identified from the 550k panel regions based on their overlap with RefSeq coordinates. Windowed Protection Scores, defined as the number of DNA fragments completely spanning a window of 120bp centered at a given nucleotide, minus the number of fragments with an endpoint within that same window, were calculated for each nucleotide 2 Kb upstream and 2 Kb downstream of each TSS. Scores were normalized to the total number of fragments within the analyzed window. A secondary normalization was subsequently performed by subtracting a running average (calculated within ±500 bp of each position) to center the data to a 1 Kb mean of zero. For each of the 4001 positions relative to the TSS, scores were averaged across all 20,000 TSS, resulting in 4001 distinct scores.

#### 9. Cancer Vs. Control Classifier Development

The main cohort was split into a training set and an independent validation using stratified random sampling on cancer type, stage, and gender using the train_test_split function from sklearn.model_selection (v1.7.0) with random_state=500.

The training set was partitioned into 4 folds with stratification applied based on type, stage, and gender, using the StratifiedKFold function of sklearn (v1.7.0) library. Within each fold, samples from 3 of the 4 folds were used for training. The training process included region selection (described below) and synthetic expansion (described below), following which the Ge80 score values for the selected regions from this expanded sample set were used as input to the binary XGBoost (v3.0.2), a gradient-boosted tree-based ensemble classifier. The model trained on these 3 folds was applied to the remaining hold-out fold. Across all four folds, this procedure yielded one out-of-fold score per training sample. These out-of-fold predictions were used to optimize regularization hyperparameters (using Optuna python library to tune parameters such as n_estimators, max_depth, learning_rate, subsample, colsample_bytree, min_child_weigh, gamma, reg_alpha, and reg_lambda) based on predictive AUC. This results in 4 models.

The above model training procedure was repeated using 20 independent permutations (also called shuffles) of the 4-fold cross-validation, resulting in 80 trained models and 20 score values per original sample in the training set. A classification threshold was established using the 99th percentile value distribution of these scores among training controls. This threshold was used for binary class prediction and sensitivity/specificity calculations. For each sample, the training cancer score was defined as the average of its 20 corresponding scores.

Once the 80 models were finalized, they were applied to the independent validation set, yielding 80 cancer scores per validation sample. For each validation sample, the cancer score was defined as the average of its 80 scores. This averaged score was then compared to the predetermined threshold for class assignment in performance evaluation.

**10. Pre-selection of regions**

From the 551,803 regions in the Twist Human Methylome panel, 152,899 autosomal regions with at least 5 CpGs were initially retained. Considering samples from the training set, for each cancer type, Mann–Whitney U tests comparing controls versus that cancer type were used to rank regions by differential methylation. For each cancer type, the top 1,000 statistically significant regions were selected. The union of these regions across cancer types comprised 7,464 regions.

To this set, we added ~12,000 regions identified from TCGA as hypermethylated in cancers, and ~2,000 regions curated from the literature as cancer-associated hypermethylated loci, resulting in a composite set of ~17.4K regions formed the feature space for further region selection and model training, with Ge80 scores from these further selected regions used as input to XGBoost.

**11. Region Selection for Model Input**

In each fold, the samples from 3 out of 4 folds were used for training in a round-robin fashion. Considering the pre-selected regions as input, Mann-Whitney tests were performed between controls and each cancer type. For each test, the top 100 statistically significant regions were selected. The union of the statistically significant regions formed the selected regions to be used for synthetic expansion and then model training for that fold. All selected regions were autosomal only.

**12. Synthetic Expansion During Training**

To improve robustness against technical variability and to increase the diversity of patterns presented to the model during training, we applied synthetic sample augmentation to each sample in the 3 folds. For every such original training sample, 10 perturbed copies were generated by pairing it with other such samples that matched on control/cancer type, stage and gender. In each of 10 augmentation rounds, a random matching sample was selected, and a new synthetic sample was created by combining approximately 80% of the features from the index sample with 20% of the features from the matched sample. Combined with the original sample, this yielded 11 training instances per original sample. This strategy increased training set variability while preserving class-specific and stage-specific signals.

**13. Classifier Locking and Independent Validation**

All aspects of region selection, hyperparameter tuning, classifier training, and threshold determination were performed using only training data or publicly available datasets, with no access to the independent validation set during model building. These were locked before application to the independent validations set. This ensured unbiased evaluation of model performance on the independent validation set.

**14. AUC, Sensitivity & Specificity Calculation**

The area under the receiver operating characteristic curve (AUC) was calculated using the roc_auc_score function from sklearn.metrics, which assesses discrimination of the cancer score across the full range of thresholds.

Sensitivity for a given category (e.g., a cancer type, a cancer type-stage subgroup, or all cancers combined) was defined as the fraction of samples in that category with scores above the predetermined cutoff. Specificity was defined as the fraction of control samples with scores below the cutoff (i.e., true negatives). To quantify uncertainty in sensitivity and specificity estimates, 95% confidence intervals (CIs) were computed as follows:

For overall sensitivity (all cancer types combined), for individual cancer type sensitivities, and for specificity, we used bootstrap resampling with 1,000 replicates. In each round, samples were resampled with replacement from the relevant group, and the statistic (sensitivity or specificity) was recomputed; the empirical distribution of bootstrap estimates was then used to derive the 95% CI.

For cancer type-stage subgroups, where sample sizes were often small, we instead used the Wilson score interval to compute 95% CIs for sensitivity and specificity. The Wilson score interval is a method for binomial proportion confidence intervals that generally provides more accurate coverage than the traditional Wald (normal approximation) interval and performs well even with small n or when proportions are near 0 or 1.

#### 15. Effect of Conversion Failure and Fragment-Level Rescue on Specificity

To evaluate the impact of the conversion rescue procedure on classifier performance, we assessed changes in specificity and sensitivity when the 80 trained models described above were applied to the independent validation set with and without the rescue step. Recall that the conversion rescue step filters cfDNA fragments by retaining only those with a conversion fraction ≥ 95%, i.e., fragments where at least 95% of non-CpG cytosines are converted to thymine, thereby reducing potential bias from incomplete enzymatic conversion.

For each independent validation sample, we generated cancer scores using standard processing with conversion rescue applied, and unrescued processing, where all filtered fragments (prior to the rescue filter) were used without applying the ≥ 95% conversion criterion.

We compared specificity (and sensitivity) between rescued and unrescued pipelines to quantify the effect of the conversion rescue step on diagnostic performance. A drop in specificity when pre-rescue features were applied supports the utility of conversion rescue in reducing false positives attributable to incomplete cytosine conversion.

#### 16. Tissue of Origin Model Development & Performance Assessment

We developed a Tissue of Origin (TOO) model using a prior-fitted tabular foundation model framework, specifically TabPFN (v2.6, Prior Labs) (Hollmann et al., Nature 637, 319–326 2025), with 32 ensemble members, to discriminate among fourteen individual cancer types. As TabPFN natively supports up to ten classes, a one-vs-rest (OvR) classification strategy was employed, in which a separate binary classifier was trained for each cancer type against all others, with final class probabilities derived from the ensemble of binary outputs, and collapsed to 9 anatomically-grouped categories.

For feature selection, we identified the top 5,000 most significant genomic regions separately from Ge80 and Lt20 methylation scores for each cancer type by performing Mann–Whitney U tests comparing that cancer type against all other cancer types (taking extra care on sex chromosomes to only compare cancers and controls of the same gender). The union of these top features (de novo TOO regions) across all cancer types and cross-validation folds served as the input feature space for model training. A secondary cross-source Mann–Whitney U selection step was applied to cap the combined feature pool at 3,000 features per fold during cross-validation, and a corresponding third pass was applied at the final training stage.

Hyperparameter optimization was not required as TabPFN is a pretrained in-context learning model that performs inference without gradient-based fitting. Four-fold cross-validation within the training set, stratified by a combined key of cancer type, disease stage, and patient gender, was used to evaluate model performance and inspect confusion matrices to characterize patterns of misclassification. A fixed random seed (42) was used throughout to ensure reproducibility.

After cross-validation, the final TOO model used the entire training set as context and was used to generate predictions for samples in the independent validation set. For each validation sample, the model produced a probability distribution over all fourteen cancer types. For each of the 14 cancer types, a separate binary TabPFN classifier was trained to distinguish that cancer type from all others. The 14 raw output probabilities were then normalized by their sum to produce a final probability distribution across all classes, from which the highest-scoring class was taken as the predicted cancer type. These predicted probabilities were collapsed to 9 anatomically-grouped categories and were used to compute Top-1 and Top-2 classification accuracy metrics, reflecting the proportion of samples for which the true tissue of origin was the highest-probability class (Top-1) or among the top two highest-probability classes (Top-2). Bootstrap resampling was used to derive confidence intervals for the accuracy of both measures.

**17. Consistency with Publicly Available Methylation Datasets**

To assess the consistency of region-level methylation patterns derived from our cfDNA fragment metrics (e.g., Ge80 and Lt20) with established tissue methylation landscapes and to obtain an initial list of regions differentially methylated in cancer, we analyzed multiple publicly available methylation datasets profiled on the Illumina Infinium HumanMethylation450 BeadChip which assays approximately 485,547 CpG sites across the human genome or the HumanMethylationEPIC BeadChip (EPIC) covering over 850,000 CpGs. Specifically, we obtained data from The Cancer Genome Atlas (TCGA), supplemented with data from the Epigenome Wide Association Studies (EWAS) consortium, and from Gene Expression Omnibus (GEO). Samples counts by source and cancer type appear in Supplementary Table 6.

From these datasets, we identified differentially methylated seed probes for each cancer type as the starting point for defining differentially methylated regions (DMRs). This was done by evaluating the following differentials probe-wise: between a given cancer type vs. (not necessarily paired) adjacent normals for that type (Specific Cancer), between a given cancer type vs. pooled adjacent normals for all types (Pooled Adjacent Normals), between a given cancer type vs. blood (Healthy Blood), and between a given cancer type vs. healthy tissue for the same type (Healthy Tissue). For the CESC and OV cancer types, where adjacent normals were not available, healthy tissue samples from GEO that were run on the HumanMethylation450 BeadChip and were available for SeSAMe normalization (employed by all samples in TCGA), were used for the Specific Cancer differential. For the STAD cancer type, since the available adjacent normals were few in number, they were combined with healthy tissue samples from GEO for the Specific Cancer differential. Samples involved in each comparison by source and cancer type appear in Supplementary Table 6.

Those HumanMethylation450 BeadChip probes that passed the differential methylation cutoffs in all comparisons comprised our set of seed probes. The cutoff strategy in each case was a function of the log fold change and the KS-test p-value as outlined in Supplementary Table 6; the specific cutoffs by cancer type are also described in Supplementary Table 6. These seed probes were expanded by a 2.5Kb window on either side to obtain an expanded list of probes.

β values for this expanded list of probes were obtained from preprocessed parquet files. Missing β values were imputed using k-nearest neighbour imputation as implemented in ChAMP (v2.22), after excluding any probes or samples with greater than 50% missingness (ProbeCutoff = 0.5; SampleCutoff = 0.5).

Region-level differential methylation analysis of β values was conducted using the linear modeling framework in DMRcate (v2.26), with sample group (cancer vs. healthy) specified as the primary explanatory variable. Probe-level associations were adjusted for multiple testing using the Benjamini–Hochberg method, with a false discovery rate threshold of 0.05, to obtain a list of statistically significant probes.

DMRs were defined by aggregating these significant probes based on spatially proximity as well as consistent differential methylation signal, applying a maximum inter-CpG distance of 500 bp (lambda = 500), a smoothing scaling factor of 5 (C = 5), a minimum of three CpG sites per region (min.cpgs = 3), and a minimum region length of 300 bp (min.length = 300). DMRs were then mapped to the overlapping target regions in the Twist Human Methylome 550K Panel in hg38 coordinates using bedtools to identify base-pair-level overlaps. Overlapping Twist panel regions were then annotated for CpG content.

To evaluate cfDNA methylation levels in our cohort for these DMRs, we retained only those Twist Human Methylome regions which mapped to a TCGA DMR and contained ≥3 CpGs. Ge80 and Lt20 scores were averaged over these regions for each sample in our main cohort, and the difference of these averaged Ge80/Lt20 score distributions between cases and controls was assessed using the Wilcoxon test.

Conversely, to evaluate methylation levels in TCGA tissue samples relative to adjacent normals or healthy tissue for differentially methylated regions in our cfDNA cohort, we mapped these regions to the Illumina Infinium HumanMethylation450 BeadChip probe coordinates (hg38) using bedtools intersectBed. For each such region, β methylation values were averaged over the overlapping probes. These region-wise β values were then averaged over all these regions and the difference of these averaged β values distributions between cancer tissue and adjacent normal/healthy tissue was assessed using the Wilcoxon test.

**18. Comparison with Previous MCED Studies**

Cancer-wise stage-wise sample counts and true positive counts were extracted from Supplementary Data of other published works for comparison and consolidated with the corresponding numbers from this study.

To enable fair comparison across studies while accounting for differences in cancer-type composition, we used a regression-based standardization framework. Stage-specific sensitivity was estimated using a binomial generalized linear model (statsmodels 0.14.6 in Python) including fixed effects for study, stage, and cancer type, together with a study-by-stage interaction term (true positive fraction ~ study*stage + cancer_type), fit using frequency weights corresponding to sample counts and family=sm.families.Binomial().

Marginal standardized stage-specific sensitivities for each study were obtained by averaging model-predicted sensitivities over the pooled cancer-type distribution observed across all studies, thereby removing confounding due to differences in cancer-type mix. Uncertainty and statistical significance of stage-specific sensitivity differences between studies were assessed using nonparametric bootstrap resampling (n=1000). One-sided bootstrap p-values were computed as the proportion of bootstrap replicates in which the estimated sensitivity difference between our study minus the previously published study was less than or equal to zero, corresponding to a directional test of superiority.

**19. Association between cfDNA Concentration and Sensitivity/TOO Accuracy**

To study the dependence of classifier sensitivity on tumor size and cfDNA extraction concentration, sensitivity was estimated using rolling windows across tumor size and cfDNA concentration distributions and stratified by disease stage.

To evaluate whether the association between cfDNA concentration and detection sensitivity was independent of cancer type and disease stage, a logistic regression model (statsmodels 0.14.6 in Python, function logit) was fitted with detection (positive classifier call) as the binary outcome. The model included cancer type and stage as categorical covariates, and cfDNA concentration as a continuous predictor modelled using natural cubic splines with four degrees of freedom to allow for non-linear dose–response relationships. The overall significance of the concentration effect, after adjustment for cancer type and stage, was assessed using a Wald test jointly applied to all spline basis terms. An additional model with further adjustment for age, gender, BMI, tobacco use, alcohol use, plasma storage time, sequencer platform, input DNA amount, and average read coverage was also evaluated for significance.

To assess whether the relationship between cfDNA concentration and detection sensitivity differed across disease stages, an exploratory interaction model incorporating stage-specific linear concentration interaction terms (stage × concentration) was additionally fitted. A linear interaction specification was used because spline-based interaction models resulted in unstable and rank-deficient fits. All analyses were performed using the statsmodels logistic regression implementation in Python without regularisation.

A similar process as the above for sensitivity was repeated for TOO accuracy.

**20. Approaches for Training-Validation Splitting**

To rigorously assess the robustness and generalizability of our predictive model under real-world heterogeneity, we partitioned the main cohort into non-overlapping training and independent validation subsets using three additional splitting schemes: site-wise, collection time-wise, and sequencing batch-wise. The division threshold in these splits was chosen to maintain the training-independent validation sample ratio as close to 60:40 as possible while balancing representation in training and independent validation sets as much as possible.

For the random split, cancer samples were divided into a training/independent validation split using stratified sampling to preserve key covariate distributions. Stratification was performed on cancer type, stage, gender, and flow cell status using the train_test_split function from sklearn.model_selection with random_state=500. This approach ensured balanced representation of major demographic and technical variables in both subsets, thereby reducing sampling bias.

The site-wise split was defined by the institution at which the sample was collected: cancer samples from sites 1–20 were assigned to the training set, and samples from sites 21–43 were assigned to the independent validation set. Site identifiers reflect the order in which sites were onboarded for sample collection, which was completed prior to laboratory processing and therefore independent of the analytical batch.

The collection-time-wise split used the temporal order of sample acquisition to simulate a prospective deployment scenario: cancer samples collected from 30 June 2022 to 31 July 2023 were used for training, and samples collected from 01 August 2023 to 04 February 2025 were used for independent validation.

The sequencing batch-wise split used chronological sequencing runs to account for potential batch effects: cancer samples sequenced between 18 August 2023 and 03 May 2024 were assigned to the training set, and samples sequenced between 21 May 2024 and 17 October 2025 were assigned to the independent validation set.

Control samples were collected from three distinct sites across India. Because of their smaller number, applying the same site-wise, collection-time-wise, or sequencing batch-wise partitions used for the cancer cohort would have introduced imbalance in key demographic and technical covariates. Therefore, we tried two approaches. First, control samples were partitioned using a stratified random split within each scheme, preserving broad demographic and technical representativeness of the healthy cohort in both training and independent validation sets. Second, control samples were partitioned as dictated by the splitting strategy (or a slight variant thereof for the site-wise split, where control samples from site 27 (n=285) were used in the training set, and control samples from sites 26 and 35 (combined n=165) were used in the independent validation set).

This combination of splitting strategies enabled thorough evaluation of model performance under potential sources of distributional shift. Consistent performance across all splits is indicative of greater robustness and generalizability in diverse real-world settings.

**21. Confounding by Demographic and Technical Covariates**

We evaluated the association between the derived classifier score and cancer outcome for each of the splitting strategies (random, site-wise, sequencing batch-wise, and collection date-wise), using logistic regression analyses with and without adjustment for potential confounders. For each split strategy, the dependent variable was cancer status (case/control), and the primary independent variable of interest was the classifier score. Covariates included age, gender, BMI, tobacco usage, alcohol usage, plasma storage time, and flow cell.

Unadjusted associations were estimated using logistic regression (using statsmodels.api.GLM with family=sm.families.Binomial(), statsmodels version 0.14.6 in Python), and adjusted associations were estimated by including the covariates listed above in the model. For each model, we derived odds ratios (ORs), 95% confidence intervals (CIs), and two-sided p-values for the cancer score to assess the strength and significance of its association with cancer status under each splitting scheme.

Additionally, we assessed the influence of individual covariates using an inverse probability weighting framework (IPW). For this purpose, we considered each of the following covariates in turn: age, BMI (dichotomized at the mean), gender, tobacco usage, alcohol usage, plasma storage time (dichotomized at the mean), and flow cell. Each covariate was considered in turn as an index variable and binarized for interpretability and weights stability. Propensity scores were estimated for the index covariate conditional on all remaining covariates (using logit from statsmodels.api, statsmodels 0.14.6 in Python), and inverse probability weights were applied to balance the distribution of these remaining covariates between levels of the index covariate, as assessed by standardized mean differences. Under successful balance, this approach yielded a marginal (population-averaged) estimate of the association between the index covariate and the outcome, independent of confounding by the other measured covariates. We estimated both weighted and unweighted logistic regression models (using statsmodels.api.GLM with family=sm.families.Binomial(), statsmodels version 0.14.6 in Python) to obtain ORs, 95% CIs, and p-values for the classifier scores.

For stratified analysis of specificity and sensitivity by tobacco usage, each of these metrics was evaluated at different training specificity thresholds and 95% confidence intervals were derived using WIlson's method.

**22. Functional analysis of the classifier regions**

**Reference annotation and background pool**

Each of the 551,803 regions was annotated with region length in base pairs (bp), CpG dinucleotide density (CpGs per 100 bp), log-normalised control depth, and ChIPseeker-derived genomic annotation class. For each region i, the log-normalised control depth was defined as log(1+mean_j(N_ij)): N_ij = C_ij × $10^9$/(L_i × D_j), where C_ij is the raw fragment count for region i in sample j, L_i is region length in base pairs, and D_j is the total mapped fragment count for sample j summed across all profiled regions. A background pool of regions was defined as a subset of the 551,803 regions, excluding the classifier regions, with ≥5 CpGs each (n = 146,722).

**Matched background region generation**

Ten matched background sets of 5000 regions each were generated by three-dimensional stratified sampling (joint distribution of region length, CpG density, and log-normalised control depth) from the background pool. Classifier regions were binned into 10 equal-frequency bins along each dimension, producing up to 1,000 distinct three-dimensional strata. For each occupied stratum, a proportional number of regions was drawn from the same stratum in the background pool; strata unoccupied in the background pool were filled by uniform random sampling from the remaining strata in the background pool. Each iteration drew 5,000 regions without replacement using a fixed seed (seed = iteration × 100 + 42). The resulting ten background sets closely mirrored the classifier regions' marginal distributions of length (mean Δ

≤ 61 bp), CpG density (mean Δ ≤ 0.10 CpGs per 100 bp), and log-normalised depth (mean Δ ≤ 0.01).

**Over-representation analysis (ORA)**

Promoter-associated genes were extracted for each region set based on promoter-proximal annotations (ChIPseeker), yielding 2,268 unique genes for the classifier region set and 2,863–2,949 genes per background set. ORA was performed independently for each gene list against five databases Reactome Pathways 2024, TISSUES Curated 2025, Azimuth 2023, Epigenomics Roadmap Histone Mark ChIP-seq, and JASPAR PWM Human 2025. using the Enrichr API via gseapy (cutoff=1.0). Significance was defined by Benjamini–Hochberg-adjusted p-value < 0.05. For each term significant for the classifier region set, an odds ratio fold-change versus background was computed as OR_FC = OR_classifier_regions/mean(OR_background regions, ten iterations), with missing background terms assigned OR = 0. An empirical p-value was derived as the proportion of background sets in which the background sets' OR met or exceeded the classifier regions' OR. Terms were classified by background recurrence: case-specific (significant in 0/10 background iterations), rarely enriched (<5/10), or background-confounded (≥5/10). Figures present the top 15 terms by OR_FC from Reactome 2024 and Epigenomics Roadmap, restricted to p_adj < 0.05.

**Cohen's d effect-size analysis**

Cohen's d was computed per region for both Ge80 methylation signal and normalized fragment depth to quantify case–control discrimination as a standardized effect size. Ge80 is defined as the proportion of reads at a region in which ≥80% of covered CpG sites are methylated. Cohen's $d = (\mu_{case} - \mu_{ctrl}) / s_{pooled}$, where $s_{pooled} = \sqrt{[((n_{case} - 1)\sigma^2_{case} + (n_{ctrl} - 1)\sigma^2_{ctrl}) / (n_{case} + n_{ctrl} - 2)]}$. Regions with pooled standard deviation < $10^{-4}$ were assigned Cohen's d = 0. A positive value for Ge80 Cohen's d indicates case-enriched hypermethylation; a negative value for depth Cohen's d indicates reduced fragment coverage in cases. Cohen's d for Ge80 methylation was plotted against Cohen's d for normalized depth, for classifier regions and for background set regions with non-0 effect sizes in both metrics (n = 49,187). Distributions of each metric were individually compared between classifier regions and the background regions sets.
